# Continuous population-level monitoring of SARS-CoV-2 seroprevalence in a large metropolitan region

**DOI:** 10.1101/2020.05.31.20118554

**Authors:** Marc Emmenegger, Elena De Cecco, David Lamparter, Raphaël P. B. Jacquat, Julien Riou, Dominik Menges, Tala Ballouz, Daniel Ebner, Mathias M. Schneider, Itzel Condado Morales, Berre Doğançay, Jingjing Guo, Anne Wiedmer, Julie Domange, Marigona Imeri, Rita Moos, Chryssa Zografou, Leyla Batkitar, Lidia Madrigal, Dezirae Schneider, Chiara Trevisan, Andres Gonzalez-Guerra, Alessandra Carrella, Irina L. Dubach, Catherine K. Xu, Georg Meisl, Vasilis Kosmoliaptsis, Tomas Malinauskas, Nicola Burgess-Brown, Ray Owens, Stephanie Hatch, Juthathip Mongkolsapaya, Gavin R. Screaton, Katharina Schubert, John D. Huck, Feimei Liu, Florence Pojer, Kelvin Lau, David Hacker, Elsbeth Probst-Müller, Carlo Cervia, Jakob Nilsson, Onur Boyman, Lanja Saleh, Katharina Spanaus, Arnold von Eckardstein, Dominik J. Schaer, Nenad Ban, Ching-Ju Tsai, Jacopo Marino, Gebhard F. X. Schertler, Nadine Ebert, Volker Thiel, Jochen Gottschalk, Beat M. Frey, Regina Reimann, Simone Hornemann, Aaron M. Ring, Tuomas P. J. Knowles, Milo A. Puhan, Christian L. Althaus, Ioannis Xenarios, David I. Stuart, Adriano Aguzzi

## Abstract

Effective public-health measures and vaccination campaigns against SARS-CoV-2 require granular knowledge of population-level immune responses. We developed a Tripartite Automated Blood Immunoassay (TRABI) to assess the IgG response against the ectodomain and the receptor-binding domain of the spike protein as well as the nucleocapsid protein of SARS-CoV-2. We used TRABI for continuous seromonitoring of hospital patients and healthy blood donors (n=72’222) in the canton of Zurich from December 2019 to December 2020 (pre-vaccine period). Seroprevalence peaked in May 2020 and rose again in November 2020 in both cohorts. Validations of results included antibody diffusional sizing and Western Blotting. Using an extended Susceptible-Exposed-Infectious-Removed model, we found that antibodies waned with a half-life of 75 days, whereas the cumulative incidence rose from 2.3% in June 2020 to 12.2% in mid-December 2020 in the population of the canton of Zurich. A follow-up health survey indicated that about 10% of patients infected with wildtype SARS-CoV-2 sustained some symptoms at least twelve months post COVID-19 and up to the timepoint of survey participation. Crucially, we found no evidence for a difference in long-term complications between those whose infection was symptomatic and those with asymptomatic acute infection. The cohort of asymptomatic SARS-CoV-2- infected subjects represents a resource for the study of chronic and possibly unexpected sequelae.

## Introduction

The severe acute respiratory syndrome coronavirus 2 (SARS-CoV-2, henceforth abbreviated as CoV2) is responsible for COVID-19 (Zhou *et al*., 2020; Zhu *et al*., 2020) and has caused millions of deaths. It has also indirectly caused many more fatalities by hijacking healthcare resources, thereby making them unavailable to patients suffering from other diseases. In addition, COVID- 19 has created profound economic distress for most travel-related industries, and has disrupted a plethora of industrial supply chains, resulting in a massive worldwide economic crisis that may cost many more human lives.

The canton of Zurich, with a population of approximately 1.5 million inhabitants, registered its first two COVID-19 cases on February 27, 2020. Zurich has seen a relatively mild first wave, with 134 deaths (and 3’785 reported cases) until June 31, 2020. However, the case numbers exploded in October, resulting in 460 deaths (and 45’516 reported cases) by December 1, 2020 (Statistisches Amt Kanton Zuerich, 2020), with hospitals working at capacity limit. In order to alleviate the direct consequences of the CoV2 pandemic, governments and public healthcare agencies need granular and reliable data on the prevalence of infection, the incidence of new infections, and the spatial-temporal oscillations of these parameters within regions of interest.

Intuitively, PCR-based diagnostics would seem suitable to fulfill the above criteria. However, practical experience has shown that this is not the case. The acquisition of representative diagnostic material for PCR has proven challenging, with deep nasal swabs being difficult to perform, uncomfortable for patients and potentially hazardous for medical personnel. Accordingly, the sensitivity of PCR diagnostics is often disappointing, with reported false-negative rates of 25% even under the best conditions (Kucirka *et al*., 2020).

Serological assays, on the other hand, address the adaptive immune responses of the host which are fundamental to limiting viral spread within individuals and populations. While they lag behind the viral infection, they can serve as both powerful epidemiological tools as well as useful clinical aids. Firstly, antibodies can be easily retrieved from many biological fluids, notably including venous and capillary blood. Secondly, antibodies typically persist for several months whereas the viral load in the upper respiratory tract frequently wanes within weeks (He *et al*., 2020). Importantly, immunological assays can be largely automated, and are thus suitable to mass screening of extremely large cohorts.

Although large serological surveys have been carried out in several countries (Anand *et al*., 2020; Carrat *et al*., 2020; Pollán *et al*., 2020; Rudberg *et al*., 2020; Ward, Atchison, *et al*., 2020), there is a lack of continuous seroprevalence data. As waning of CoV2 antibodies has been reported in multiple instances (Buss *et al*., 2020; Ibarrondo *et al*., 2020; Q. X. Long *et al*., 2020; Seow *et al*., 2020; Ward, Cooke, *et al*., 2020; Choe *et al*., 2021), single timepoint serology estimates may yield misleading insights into the true extent of CoV2 spread. We, therefore, aimed to investigate the evolution of the CoV2 seroprevalence in the canton of Zurich, a particularly low prevalence setting during the first and second waves in 2020, using an in-house developed tripartite automated blood immunoassay (TRABI) already employed in multiple studies (Cervia *et al*., 2020; Emmenegger, Saseendran Kumar, *et al*., 2021; Emmenegger *et al*., 2022; Gehlhausen *et al*., 2022; Schneider *et al*., 2022). Continuous immunosurveys were conducted in a large cohort of the University Hospital of Zurich (n=55’814 samples) and blood donors from the Blood Donation Services of the canton of Zurich (n=16’291), over a period from December 2019 to December 2020, i.e. prior to the onset of the vaccination campaigns. Apart from assessing the underlying cumulative incidence, we aimed to build a foundation for the subsequent identification of sequelae in the clinically well characterized hospital patients. To this end, we have made use of available ICD-10 codes and free-text reports to elucidate whether seropositivity is associated with disease entities beyond those already reported. Finally, we invited serologically tested hospital patients to participate in an online health survey to investigate the health status of seropositive patients post COVID-19, with the first infection dating back more than 500 days (median). These combined seroepidemiological and nosoepidemiological endeavors, together with the close monitoring of ongoing vaccination efforts and variants of concerns (VOCs), are likely pivotal in enhancing our understanding on how to manage the current as well as future pandemic outbreaks.

## Results

### TRABI: a miniaturized high-throughput ELISA for multiple CoV2 antigens

Here we assessed the changes in CoV2 seroprevalence in the population of the canton of Zurich (n=1.5 million) between December 2019 and December 2020. To this end, we developed a tripartite automated blood immunoassay (TRABI) utilizing contactless acoustic dispensing (Kulesskiy *et al*., 2016; Kryštůfek and Šácha, 2019) to transfer diluted plasma droplets (2.5 nl) into high-density 1536-well plates (total volume: 3 µl) and measuring the IgG response against viral proteins by immunocolorimetry (**Fig. 1A** and **Fig. S1A** for detailed procedure).

**Fig. 1.**
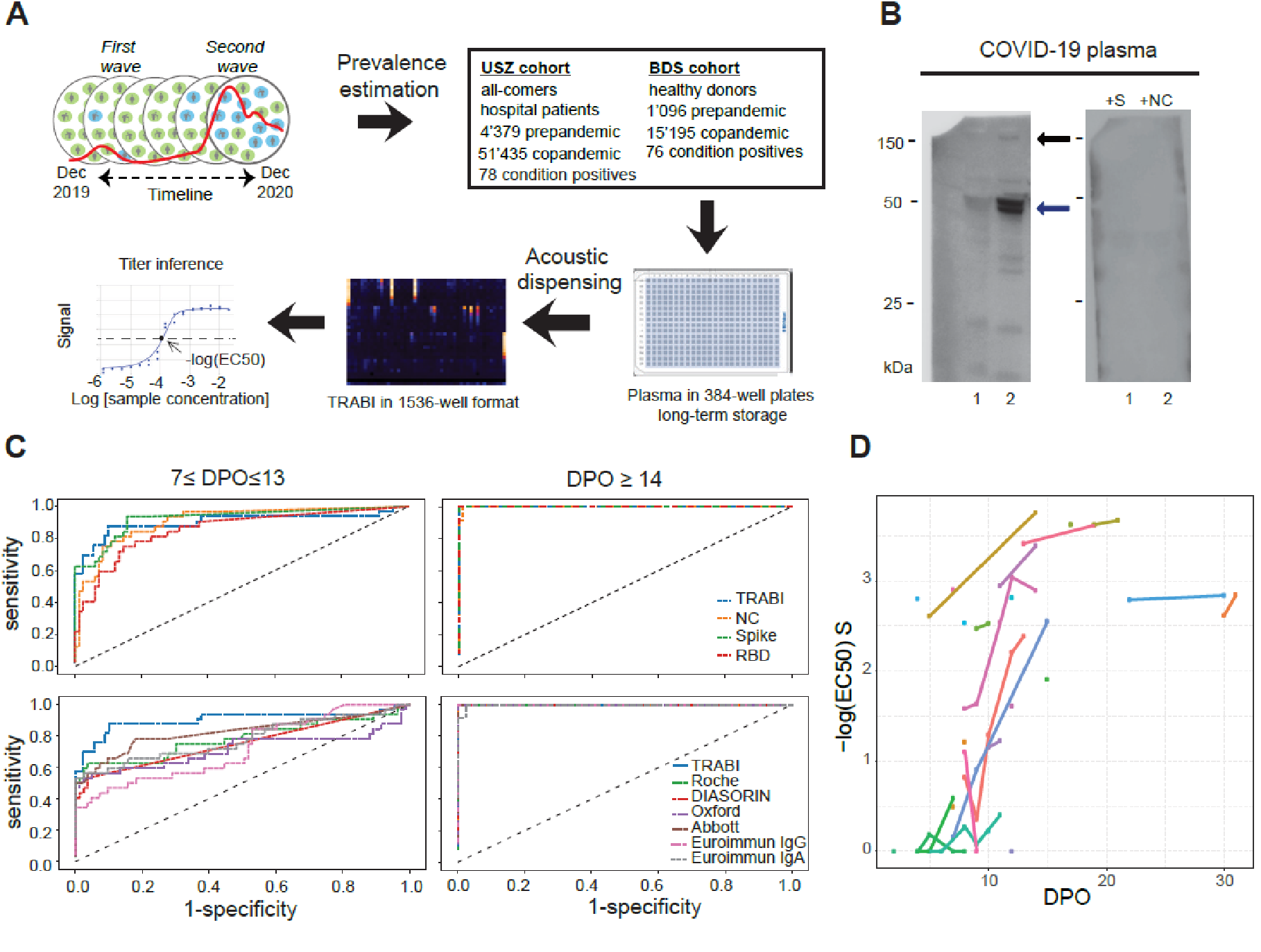
Study overview and establishment of serological pipeline. **A**. To estimate the prevalence of CoV2 seropositivity in the population, prepandemic and copandemic samples from two independent cohorts were analyzed by high-throughput microELISA (TRABI). IgG titers against S, RBD and NC were determined and the -log(EC_50_) was inferred by regression analysis. **B**. Vero cells infected with CoV2 (lane 2), but not uninfected cells (lane 1), showed signals corresponding to S (black arrow) and NC (blue arrow, pointing at two bands) when immunoblotted with COVID- 19 patient plasma. NC protein undergoes a proteolytic cleavage in SARS-CoV infected VeroE6 cells, resulting in two distinct bands of around 46 and 43 kDa. We confirmed the identity of the two bands by probing with an anti-NC antibody (Sino Biologicals, data not shown). Spiking of COVID-19 patient plasma with recombinant S and NC led to the disappearance of all signals. **C**. Upper panel: Using 53 samples from confirmed CoV2 patients and 83 prepandemic samples, we assessed the specificity-sensitivity relationship for all antigens individually and after combining all results into a single score (TRABI) using QDA-based posterior probability. Between 7 and 13 dpo, approximately 60% of samples were positive (posterior probability >0.5) at 100% specificity cutoff, whereas 100% sensitivity was reached at ≥ 14 dpo. Lower panel: COVID and prepandemic samples were used to assess the performance of TRABI, commercial tests (Roche, DiaSorin, Abbott, Euroimmun) and an assay developed at the Target Discovery Institute (Oxford). While all tests scored equally at ≥14 dpo, TRABI outperformed all other assays at ≤13 dpo. **D**. Time course of IgG response in 55 samples from 27 COVID patients. IgG antibodies were reliably detectable at ≥13 dpo. Colors represent individual patients.

In order to identify the most suitable viral targets for TRABI, we infected Vero cells with wild-type CoV2 virus. Cell lysates were then subjected to Western blotting using the plasma of patients with confirmed COVID-19 (n=7). The bands corresponding to the S and NC proteins were prominently visible in infected cells, but were undetectable in non-infected cells and were suppressed by adding soluble S and NC antigen to the patient plasma before incubation with the Western blot (**Fig. 1B**). Accordingly, we selected the CoV2 spike protein (Song *et al*., 2018), the receptor binding domain (RBD, amino acids 330-532 of the S protein), and the nucleocapsid protein (NC, amino acids 1-419) as target antigens for TRABI. Each sample was tested at eight consecutive two-fold dilution points (1:50 to 1:6’000), and the resulting data were fitted to a sigmoidal curve by logistic regression. The inflection point (or –log_10_(EC_50_)) of each sigmoid was defined as the respective antibody titer.

As reference samples for assay establishment, we utilized a collective of 55 venous plasma samples drawn at various days post onset of symptoms (dpo) from 27 RT-qPCR confirmed patients suffering from COVID-19 and hospitalized at the University Hospital of Zurich (USZ, true positives, see **Table 1** and **Table S1**), as well as 90 anonymized USZ samples from the prepandemic era (true negatives). We then constructed receiver-operating-characteristics (ROC) curves to assess the assay quality for each antigen individually. Finally, we created a composite metric that integrates S/RBD/NC measurements using quadratic discriminant analysis (QDA). While each single antigen showed excellent discrimination of negatives and positives on samples drawn at ≥14 dpo, the compound models outperformed the individual antigen measurements at 7-13 dpo, where the emergence of an IgG response is expected to be variable (**Fig. 1C**, upper panel). We therefore used the QDA modeling assumptions to infer the prevalence in large cohorts based on the distributional information of true negatives and true positives using information gained from all three antigens.

**Table 1.** Cohort characterization on age and sex.

|  |  | Assay establishment | USZ cohort | BDS cohort | All cohorts |
| --- | --- | --- | --- | --- | --- |
| *Total* | Individuals, number | 117 | 55814 | 16291 | 72222 |
|  | Median age (IQR), years | 62 (52-70) | 55 (40-68) | 42 (28-54) |  |
|  | Sex, female % | 37 | 47 | 41 |  |
|  | Sex, male % | 63 | 53 | 59 |  |
| *Copandemic* | Individuals, number | 27 | 51435 | 15195 | 66657 |
|  | Median age (IQR), years | 62 (52-70) | 55 (40-68) | 42 (28-54) |  |
|  | Sex, female % | 37 | 47 | 41 |  |
|  | Sex, male % | 63 | 53 | 59 |  |
| *Prepandemic* | Individuals, number | 90 | 4379 | 1096 | 5565 |
|  | Median age (IQR), years | / | 54 (39-68) | / |  |
|  | Sex, female % | / | 48 | / |  |
|  | Sex, male % | / | 52 | / |  |

To benchmark TRABI, we compared the results with a high-throughput assay – at the time of testing still under development – at the University of Oxford as well as assays commercialized by Roche (Elecsys), DiaSorin, EuroImmun, and Abbott (**Fig. 1C**, lower panel). This comparative assessment was based on 136 of 146 samples (10 samples were removed from the analysis because of insufficient sample volume to perform all tests). While all assays displayed 100% specificity/sensitivity at late time points, TRABI scored best at early time points, also when additionally compared to a lateral-flow assay (**Fig. S2A**). When these results were plotted as a function of dpo, a temporal pattern emerged consistent with the gradual emergence of IgG antibodies within 14 dpo (**Fig. 1D**).

### Characterization of cohort used for seroprevalence estimates from December 2019 to December 2020

Anti-CoV2 antibodies were measured with TRABI in 66’630 copandemic samples (collected between December 2019 and December 2020), 51’435 belonging to patients of the USZ and 15’195 to blood donors. The median age of the USZ patients was 55 (40-68) years (**Table 1** and **Fig. S3A**) and 42 (28-54) years of the blood donors (**Table 1** and **Fig. S3B**), which was stable over the time span of our measurements for the USZ patients (**Fig. S3C**) but showed deviations for the blood donors, with a decrease in overall age between April and August, 2020, followed by an increase in age from henceforth (**Fig. S3D**). The sex distribution in the USZ sample was stable over time, with a female/male ratio close to parity (**Fig. S3E**). The BDS sample contained slightly more men than expected (**Fig. S3F**). Most of the hospital patients included in this study were adult residents of the canton of Zurich (**Fig. S4A**) and were treated in one of the many clinical departments (**Fig. S4B**), the highest number in Medical Oncology and Hematology, followed by Cardiology, Infectious Diseases and Hospital Hygiene, Rheumatology, and Gastroenterology and Hepatology. The distribution of samples originating from these hospital wards was relatively stable over time (**Fig. S4C**). 5’345 distinct ICD-10 codes were assigned to hospital patients, of which the 50 most common ones are summarized in **Table S2**. Within these 50 ICD-10 codes are many of the common diseases like ‘essential primary hypertension’ (ICD-10: I10.00), ‘type II diabetes mellitus’ (ICD-10: E11.9), or ‘heart failure’ (ICD-10: I50) but also ‘chronic kidney disease’ (ICD- 10: N18), and ‘malignant melanoma of skin’ (ICD-10: C43).

### Temporal evolution of the CoV2 epidemic in the greater area of Zurich

5’475 prepandemic samples collected before December 2019 were used as condition negatives (see **Table 1**) and 154 copandemic samples identified as condition positives. Their annotation as condition positives was performed post-hoc using USZ and BDS databases in the absence of serological data. First, we identified all USZ samples with known positive CoV2 RT-qPCR results (n=320). Condition-positive samples (n=78) were defined as those with (1) clinically manifest COVID-19 pneumonia and (2) positive RT-qPCR for CoV2 and (3) venipuncture occurring ≥ 14 days after the positive qPCR to account for seroconversion. To avail of condition positives from the cohort of blood donors, 76 samples from convalescent individuals with PCR-confirmed CoV2 infection recruited for a plasmapheresis study were included. In addition to the QDA-based model that assumes that both the condition-positive and negative data follow distinct multivariate Gaussian distribution with unequal covariances (**Fig. 2A, B**), we tested a model based on Gaussian distributions with equal covariances: linear discriminant analysis (LDA) (**Fig. S5A, B**). LDA allows to verify the distributional assumptions more readily (**Fig. S5C, D**). Using the distributions of the condition negatives and the condition positives, we computed the posterior probability (i.e. the probability of an individual to be seropositive as modeled via the distribution of the known condition-negatives and known condition-positives) for all data points. The respective ROC curves were then plotted (**Fig. S5E, F**). At 100% specificity, we identified 78% of the annotated true positives for the USZ (**Fig. S5E**) and 67% annotated true positives for the BDS cohort (**Fig. S5F**). For both the USZ and the BDS cohorts, the sensitivity increased rapidly with a slight decrease in specificity (at a false-positive-rate of 0.001, we identified 82% condition positives for USZ and 89% for BDS).

**Fig. 2.**
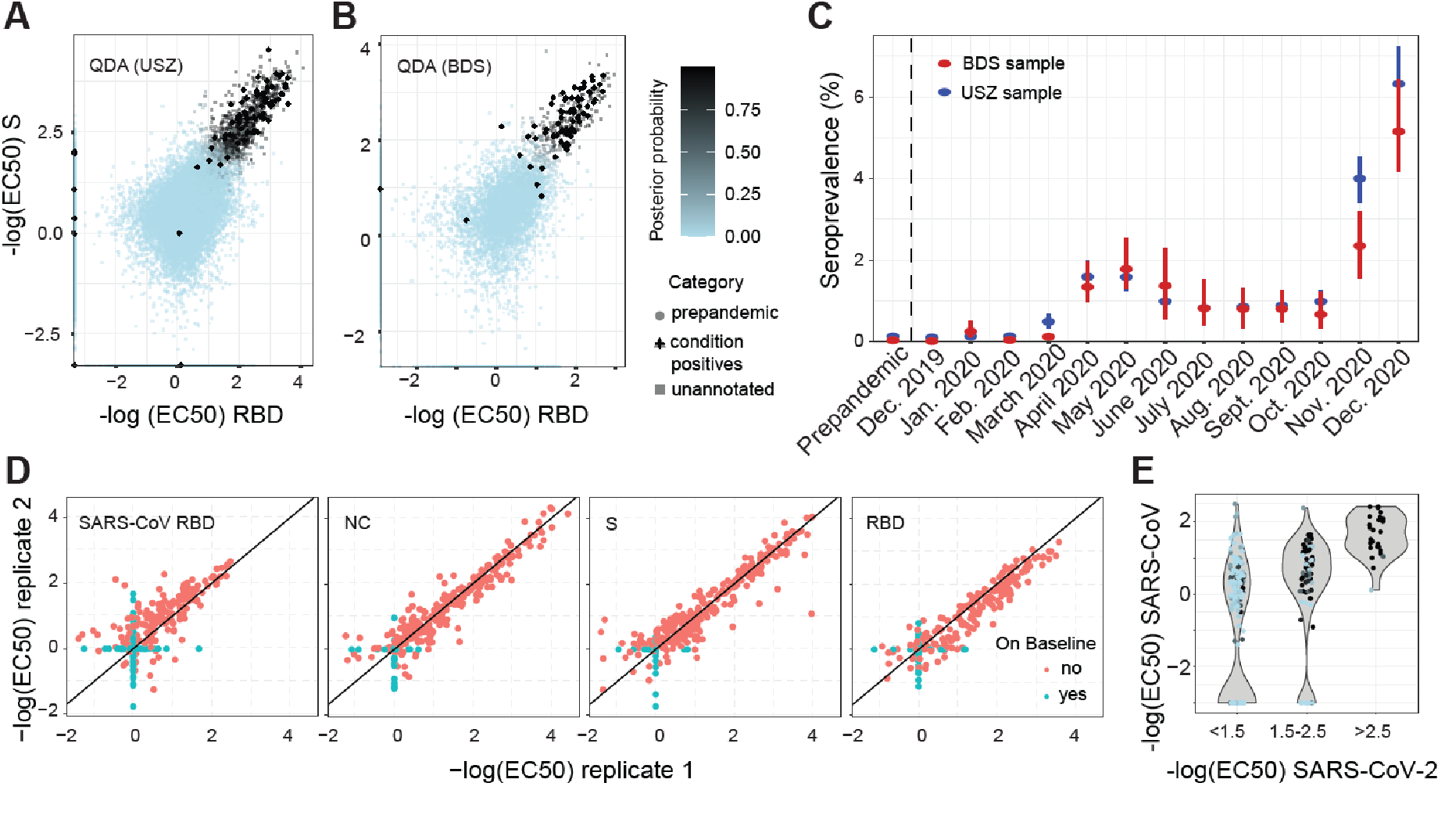
Evolution of CoV2 prevalence in a cohort of Zurich University Hospital (USZ) patients and donors from the blood donation service (BDS). **A-B**. Inflection points of dilution curves, denoted -log(EC_50_), of plasma titrated against S and RBD in the USZ and BDS cohorts. Posterior probabilities were calculated using QDA assuming a multivariate Gaussian distribution. **C**. Prevalence of CoV2 seropositivity in prepandemic (before December 2019) and co-pandemic samples (from December 2019 to December 2020) estimated using the posterior probabilities from the multivariate Gaussian distribution (QDA). Bar: 95% confidence intervals (CI). **D**. TRABI reproducibility was assessed using duplicates run in pairs of independent assay plates. **E**. To assess potential cross-reactivity of CoV2 seropositive individuals, we tested 200 high-scoring samples and 112 random samples for binding to the RBD of SARS-CoV. CoV2 RBD binders with a high posterior probability (same color maps as in B) segregated within the higher anti-SARS- CoV-RBD titers.

We then applied the QDA-based probability model to estimate the monthly prevalence, from December 2019 to December 2020, using the USZ and the BDS cohorts. No substantial shift above baseline was inferred for samples screened until February 2020 (**Fig. 2C**). In March 2020, the USZ-based prevalence increased to 0.5% (95% confidence intervals: 0.3%-0.7%) and to 1.6% (CI95%: 1.2%-2.0%) in April 2020, with blood donors displaying a comparable course of seroconversion, with the prevalence approximating 1.3% in April (CI95%: 1.0%-2.0%). The blood donors then reached a first peak in May 2020, with a prevalence of 1.8% (CI95%: 1.3%-2.5%), while the USZ patients plateaued. Following an initial decline in June (USZ: 1.0% (CI95%: 0.8%-1.2%), BDS: 1.4% (CI95%: 0.6%-2.3%)), the seroprevalence fluctuated at around 0.8% over the course of the summer. These summer months were generally characterized by a low reported incidence (4,106 new PCR-confirmed cases and 16 COVID-19-associated deaths from July 1 to September 30 in the canton of Zurich (Statistisches Amt, 2020)), until a second wave surged in October. A sharp rise in seroprevalence was observed for November (USZ: 4.0% (CI95%: 3.4%-4.5%), BDS: 2.4% (CI95%: 1.5%-3.2%)) and beginning/mid-December 2020 (USZ: 6.3 (CI95%: 5.5%-7.2%), BDS: 5.1% (CI95%: 4.2%-6.4%)).

To assess the technical reproducibility of TRABI, we repeated the assay on 200 and 112 randomly selected positive and negative samples, respectively. This repeat screen was found to reproduce the original TRABI results (R^2^ = 0.85, **Fig. 2D** and **Fig. S6**).

Antibodies against the RBD of SARS-CoV can bind to the CoV2 RBD (Tai *et al*., 2020). We therefore tested whether samples with high anti-CoV2-RBD titers display cross-reactivity with SARS-CoV RBD. For visualization, we binned samples into groups of absent, moderate and high CoV2 RBD titers (–log[EC_50_] < 1.5, 1.5-2, and > 2.5, respectively) and computed their respective QDA- derived posterior probability. For individuals with CoV2 RBD titers < 2, a small fraction showed binding to SARS-CoV RBD at –log(EC_50_) > 2 (**Fig. 2E**). However, those with strong binding properties to CoV2 RBD (> 2.5) clustered at high values for SARS-CoV RBD, indicating that some anti-CoV2 RBD antibodies were cross-reactive to SARS-CoV RBD.

### Post-stratification for age and sex and removal of patients admitted because of COVID-19

We then stratified the seroprevalence data according to age and sex, for both cohorts (**Fig. S7A** and **S7B**). As the age and sex distributions of the USZ and BDS cohorts are not entirely congruent with the distributions within the general population (**Fig. S3A** and **3B**), we employed a post-stratification on sex and age using distributional information from the population of the canton of Zurich (**Fig. 3A** and **3B**). However, this correction led to only minor changes (maximal effect observed: 5.1% (CI95%: 4.2%-6.4%) unadjusted versus 4.0% (CI95%: 3.1%-5.1%) adjusted for age and sex, for blood donors in December 2020) in the calculated prevalence, suggesting that the two cohorts appropriately reflect the seroprevalence of the adult population.

**Fig. 3.**
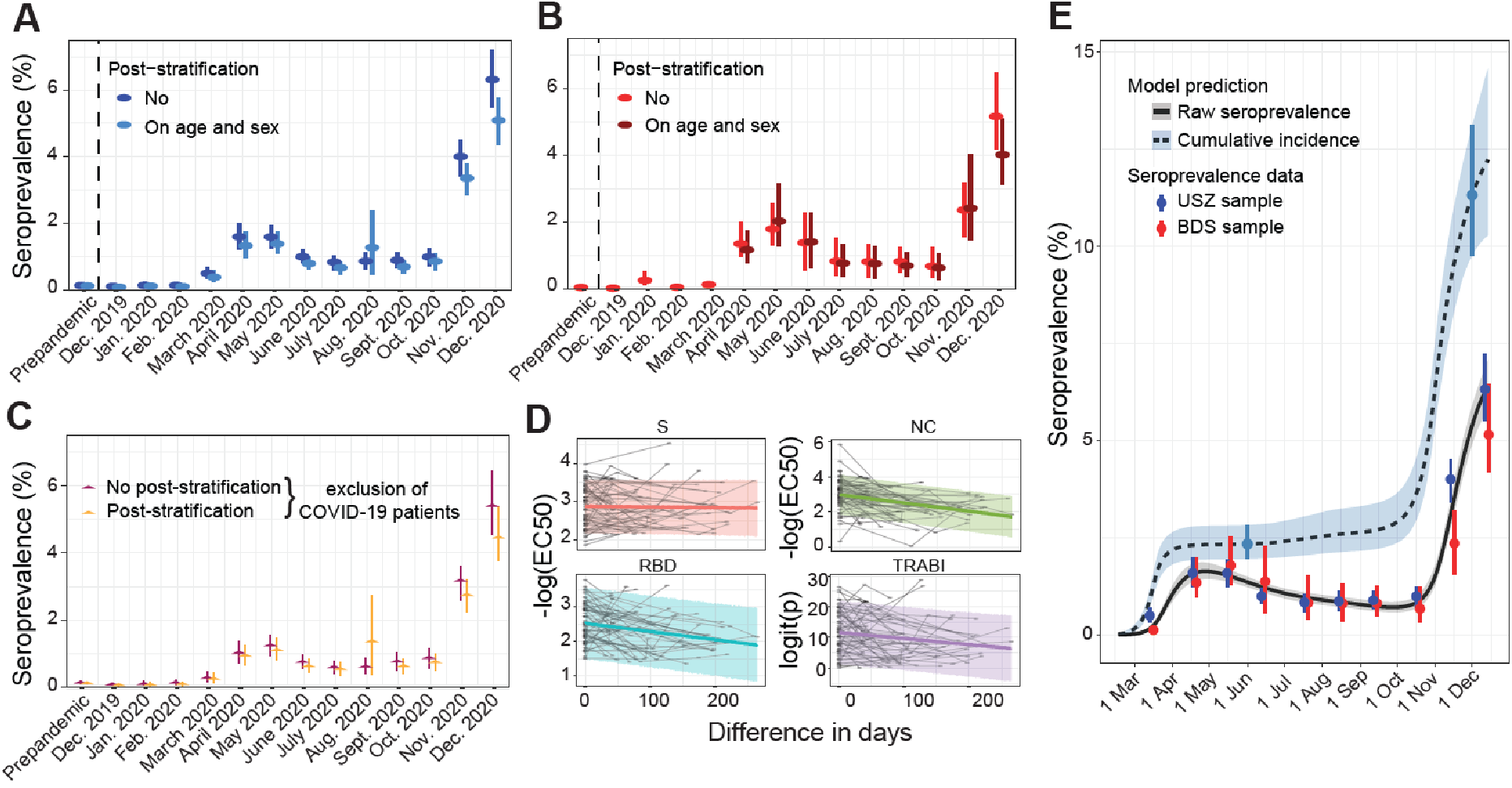
Post stratification and antibody waning. **A**. Seroprevalence in USZ patient cohort after post-stratification on age and sex using the age and sex distributions of the canton of Zurich. **B**. Seroprevalence in BDS cohort after post-stratification on age and sex using the age and sex distributions of the canton of Zurich. **C**. Seroprevalence in USZ patient cohort after removal of patients hospitalized because of COVID-19, for both raw seroprevalence and seroprevalence data after post-stratification on age and sex. **D**. Antibody waning observed with longitudinal sampling. **E**. Cumulative incidence or infection attack rate calculated from seroprevalence corrected for waning.

Additionally, we aimed to assess the extent of a bias posed by patients with severe COVID-19, hospitalized at the USZ for this reason. We thus removed patients (1) admitted to the Infectious Disease and Hospital Hygiene or the Internal Medicine wards or (2) with ICD-10 codes J96.00 (‘Acute respiratory failure’) and U.99.0 (‘Special procedures for testing for CoV2’) from the dataset and re-evaluated the course of seroprevalence for the cohort of hospital patients. We found that COVID-19 patients contribute to the prevalence observed during both the first as well as the second wave (**Fig. 3C**). Yet, the application of post-stratification on age and sex and the removal of COVID-19 patients did not change the overall dynamics of seroprevalence.

### Antibody waning and cumulative incidence

The decrease in seroprevalence observed after the peak of the first wave is suggestive of waning of antibodies on the population level. The availability of repeated samples from the hospital patients allowed us to explore the titers individually. Using data from 65 individuals with a posterior probability ≥ 0.5 and at least two seroestimates, we observe a decrease of all measurements, except for the S protein, over time, including the compound metric (**Fig. 3D**), in line with a previous report (Fenwick *et al*., 2021). We then estimated the half-life of the decrease of the antibody titer directly from the seroprevalence data, using an extension of the classic Susceptible-Exposed-Infectious-Removed (SEIR) model (Grinsztajn *et al*., 2021). Assuming an average time to seroconversion of 14 days (Guo *et al*., 2020; Jiang *et al*., 2020; Q.-X. Long *et al*., 2020), an average generation interval of 5.2 days (Ganyani *et al*., 2020) and an average time from disease onset to death of 20.2 days (Linton *et al*., 2020), the overall waning rate observed on the level of the population is 75 (CI95% 55-103) days (unadjusted) or 88 (CI95%: 61-128) days (post-stratification for age and sex), similar to what was reported by others (Menges, Zens, *et al*., 2021). We then computed the cumulative incidence of CoV2, i.e. the seroprevalence corrected for antibody waning, for the population of the canton of Zurich (**Fig. 3E**). The cumulative incidence first raised in March and slowly but gradually increased over the summer period, cumulating to 2.3% (CrI95%: 2.0%-2.8%) in June, 2020. A sharp escalation was detectable at the beginning of November, mounding in an cumulative incidence of 12.2% (CrI95%: 10.3%-14.6%)) in mid-December 2020. This suggests that over 180’000 people had contracted CoV2 until mid-December 2020 in the canton of Zurich. Thus, the cumulative number of cases detected by PCR (55’375 until 13^th^ of December 2020 (Statistisches Amt, 2020)) is likely to underestimate the true prevalence by approximately factor 3 on average. However, the hidden epidemic ratio (i.e. the number of unobserved cases for each reported case) has changed over time, with a drastic underestimation of cases at the time of the first wave, a clearly improved precision around summer 2020, and a significant underestimation during the second wave (**Fig. S7C**).

### Spatiotemporal seromonitoring in USZ patients covering two waves

We aimed to further depict the evolution of seroprevalence in the canton of Zurich. As we avail of the zip codes, we first mapped the total number of hospital patients per zip code for the months March-July (first wave) and September-December (second wave) 2020 (**Fig. 4A** and **4B**), only considering the fraction of patients from the canton of Zurich (**Fig. S4A**). We then investigated the fraction of seropositive hospital patients over the total number of hospital patients per zip code, for above time periods but restricting the analysis to municipalities with at least 50 patients in total, to avoid statistical variability. In line with the overall increased seroprevalence, we observed more than double the number of municipalities (97) showing a prevalence higher than 2% during the second wave, compared to 45 in the first wave (**Fig. 4C** and **4D**). This result is indicative of that that the epidemic outbreak in Zurich is not focal but extends throughout the canton, with similar rates of increase. The decrease of the fold-change of positive/total cases in the city of Zurich compared to the rest of the canton of Zurich from the first to the second wave (**Fig. S7D**) is substantiating the observation that after a slightly more localized first outbreak and a remission phase, the second wave is characterized by a non-focal spread.

**Fig. 4.**
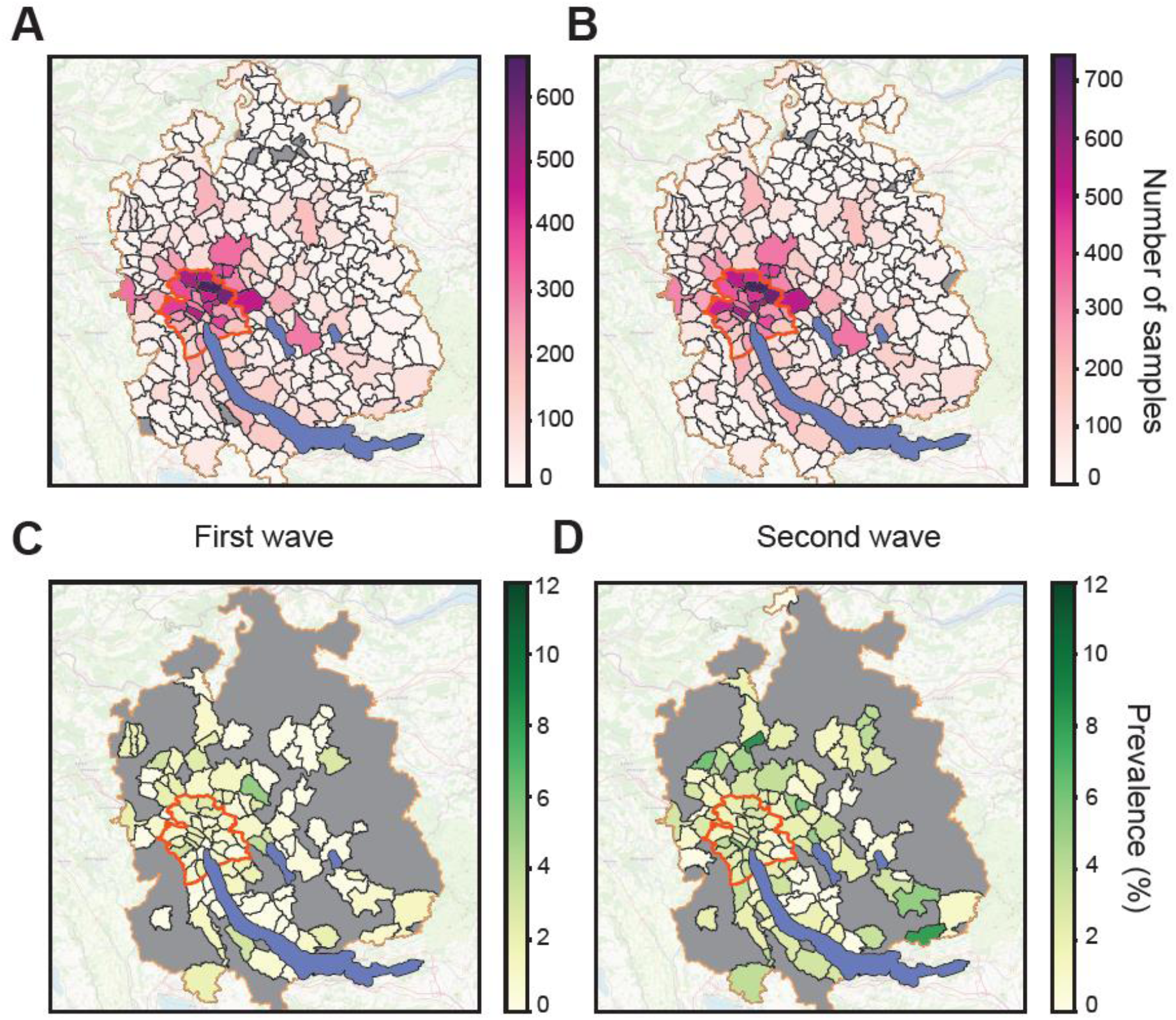
Seroprevalence maps for municipalities in the canton of Zurich. **A**. Samples of hospital patients residing in Zurich sorted according to zip codes. Data from January 2020 to June 2020, including first wave. **B**. Samples of hospital patients residing in Zurich sorted according to zip codes. Data from July 2020 to December 2020, including second wave. **C**. Seropositive samples of hospital patients residing in Zurich sorted according to zip codes. Data from January 2020 to June 2020, including first wave. **D**. Seropositive samples of hospital patients residing in Zurich sorted according to zip codes. Data from July 2020 to December 2020, including second wave. C and D: Only municipalities with at least 50 samples/zip code are displayed.

### Association with demographic and medical data

We then investigated the association between CoV2 seropositivity and disease. First, we retrieved the International Classification of Disease (ICD-10) codes entered by medical encoders of the hospital for insurance purposes, along with age and sex. Using multiple logistic regression in a Bayesian framework, we found positive associations between seropositivity and ICD-10 codes U99.0 (‘Special procedures for testing for CoV2’), J96.00 (‘Acute respiratory failure’), I48.3 (‘Typical atrial flutter’), U69.0 (‘Pneumonia acquired in the hospital, classified otherwise’), Y82.8 (‘Other medical devices associated with adverse incidents’), N17.83 (‘Other acute kidney failure’), D64.8 (Other anemia‘’), E11.91 (‘Type 2 diabetes mellitus without complications’), E87.1 (‘Hypo-osmolality and hyponatremia’), and male sex (**Fig. 5A**). However, only U99.0 and J96.00 displayed a consistently distinct positive association after regularization with horseshoe and LASSO priors. Negative associations were found with ICD-10 code Z11 (‘Special procedure to the diagnosis of infectious and parasite diseases’), while other codes did not persist after regularization and were probably spurious. Next, to better account for the hierarchically structured web of ICD-10 codes and their interdependencies, we employed a network-based representation (Jeong *et al*., 2017), aiming to investigate differentially structured nodes in ICD-10 codes, clinical departments, age, and sex, in CoV2-seropositive and seronegative USZ patients. We did not identify any distinctive motif of enriched ICD-10 codes between the seropositive and seronegative patients (**Fig. S8A**), based on topological network scores derived from the Mcode algorithm (Bader and Hogue, 2003), indicating no greatly altered disease networks as a function of a CoV2 infection. Furthermore, nonlinear Uniform Manifold Approximation and Projection for Dimension Reduction (UMAP), adjusted for binary data using a cosine metric as well as principal component analysis (PCA) did not reveal any separate cluster for seropositive patients when projecting the variability of the dataset into two-dimensional space, neither when including sex as a feature alongside ICD-10 codes (**Fig. S8B, C**) nor upon exclusion of female/male sex (**Fig. S8D, E**). The exclusion of patients without ICD-10 codes did not change this, both applying a binary (**Fig. S8F**) as well as an Euclidean distance metric (**Fig. S8G**). Lastly, in a more targeted analysis, we split our dataset into (1) seropositive COVID-19 patients hospitalized in the Infectious Diseases or Internal Medicine units (n=240), (2) seropositive patients associated with other clinical wards (n=483), and (3) randomly selected seronegative patients (n=631), aiming to interrogate the three groups for differences in potential complications of CoV2 infections recently discussed (Almqvist *et al*., 2020; Elena and Philippe, 2020; Varatharaj *et al*., 2020; Buehler *et al*., 2021; Emmenegger, Saseendran Kumar, *et al*., 2021), in ICD-10 codes as well as in free-text medical reports. As control indications, we queried for known risk factors (e.g. type II diabetes, obesity, hypertension, COPD, chronic kidney disease) for hospitalization and COVID-19 disease severity (Jehi *et al*., 2020; Petrilli *et al*., 2020; Cummins *et al*., 2021) and for well-established CoV2 complications (respiratory insufficiency, dyspnea, ARDS, pulmonary embolism, pneumonia).

**Fig. 5.**
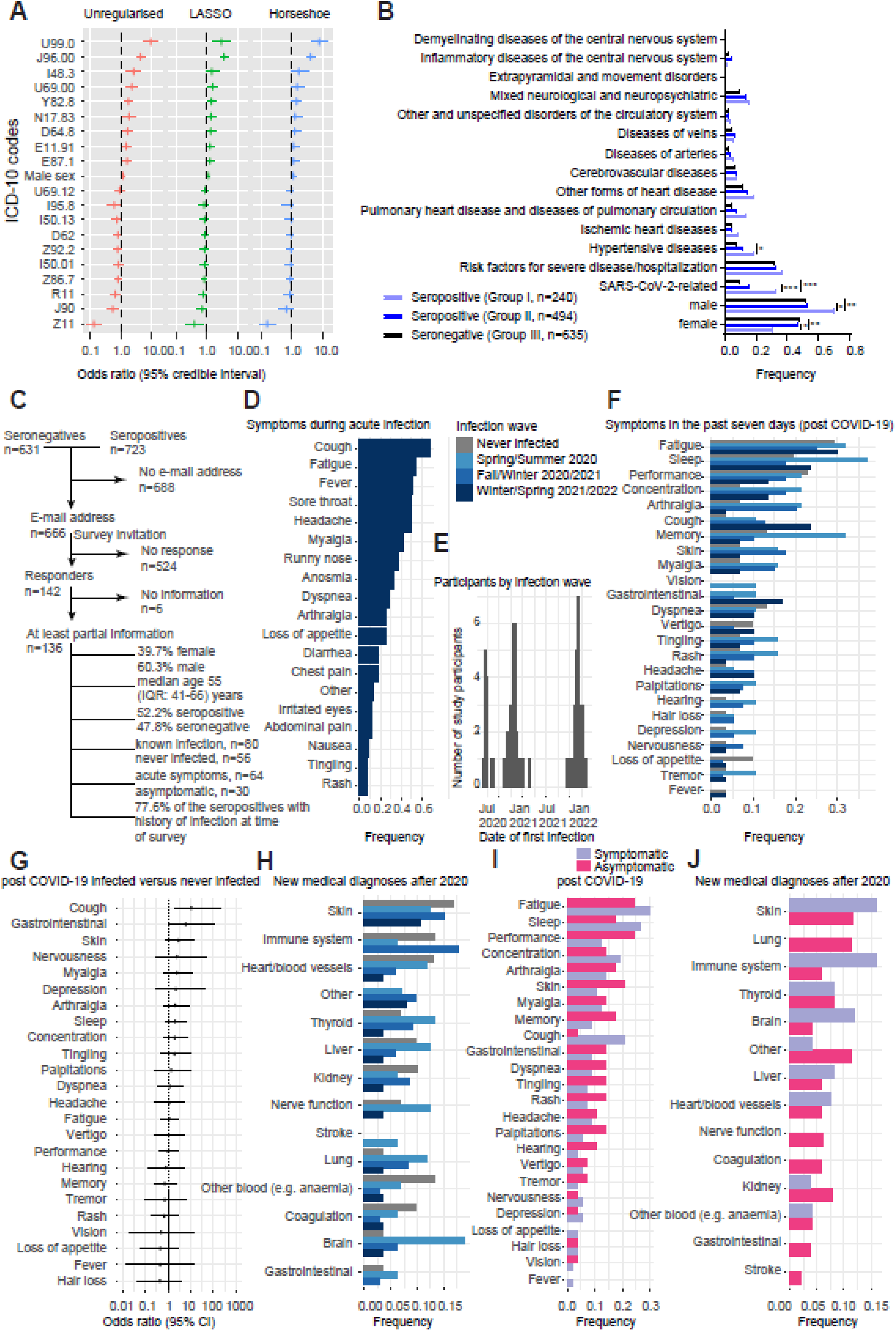
Exploratory analysis of CoV2 seropositivity with ICD-10 codes and free-text medical reports. **A**. Multiple logistic regression after logit-transforming the posterior probability in a Bayesian framework. Shown is the odds ratio with 95% credible interval. **B**. Group-wise frequencies (number of counts divided by total per group) of different disease classes/conditions. Fisher’s exact test was performed to test for deviations from expected frequencies. Male patients were much more prevalent among the seropositive COVID-19 patients (69.6% male versus 30.4% female) than in the two other groups, at statistical significance (adjusted p-values < 0.002). Hypertensive diseases were more prevalent in COVID-19 patients compared with seronegative patients (adjusted p-value = 0.002). **C**. Flowchart for the inclusion of serologically tested individuals participating in the follow-up online health survey in April/May 2022. A total of 136 individuals provided informed consent and filled the electronic questionnaire, among which 80 reported a known CoV2 infection up to questionnaire completion. **D**. Frequency of symptoms reported by online health survey participants reporting a symptomatic infection prior to April/May 2022 (n=64). **E**. Date of first infection reported by online health survey participants with a known infection prior to April/May 2022 (n=80, 2 participants with missing date). Three pandemic waves were reflected in the data: Spring/Summer 2020 (first wildtype CoV2 wave), Fall/Winter 2020/2021 (second wildtype CoV2 wave) and Winter/Spring 2021/2022 (omicron CoV2 wave). **F**. Proportion of online health survey participants reporting to have experienced within the last seven days prior to questionnaire completion, stratified by prior infection status and pandemic wave during which the infection occurred. **G**. Odds ratio of experiencing specific symptoms within the last seven days prior to questionnaire completion in the group of online health survey participants with reported known prior infection compared to the group of participants without known infection, based on multivariable logistic regression models adjusted for age and sex (central estimate: odds ratio, error bars: 95% confidence interval (95%CI). **H**. Proportion of online health survey participants reporting having received a new medical diagnosis after 2020, stratified by prior infection status and pandemic wave during which the infection occurred. **I**. Proportion of online health survey participants reporting to have experienced within the last seven days prior to questionnaire completion, stratified by symptoms during acute infection. **J**. Proportion of participants reporting having received a new medical diagnosis after 2020, stratified by symptoms during acute infection. Adjusted p-values ≤ 0.01: *. Adjusted p-values ≤ 0.001: **. Adjusted p-values ≤ 0.0001: ***.

While the three groups did not display statistically significant differences (Fisher’s exact test, p-value adjusted for multiple comparisons) in the presence of risk factors, the seropositive COVID- 19 patients (group I) differed significantly from the seropositive patients from other clinical wards (group II, adjusted p-value<0.0001) and from the seronegative patients (group III, adjusted p-value<0.0001) in known CoV2-associated diseases, illustrated in **Fig. 5B**. None of the neurological or cardiocirculatory conditions investigated showed significant differences between the groups, except for hypertensive diseases that were more prevalent in COVID-19 patients compared with seronegative patients (adjusted p-value=0.002). Age classes were slightly different in groups I compared to group II (p-value=0.0016, Mann-Whitney U test) but not in any other group-wise comparison, with a median age of 58 (IQR: 46-66) years, 53 (IQR: 37-65) years, and 54 (IQR: 39-68) years in the three groups. Male patients were much more prevalent among the seropositive COVID-19 patients (69.6% male versus 30.4% female) than in the two other groups (**Fig. 5B**; adjusted p-value < 0.002).

### Follow-up online health survey to investigate potential post COVID-19 condition

Even if patients do not experience overt COVID-19-associated pneumonia or other severe symptoms during acute infection, CoV2-infected individuals may develop post COVID-19 conditions (Kamal *et al*., 2021; Menges, Ballouz, *et al*., 2021; Yong, 2021; Tran *et al*., 2022). We invited hospital patients whose blood had been analyzed by TRABI to participate in an online health survey. In 1’354 database entries of hospitalized patients (n=723 seropositives, n=631 seronegatives), e-mail address was available for 666 allowing to send a survey invitation. Of those, 142 consented to participate and completed the questionnaire, of which 136 contained at least some information that could be used for analysis (participation rate 10.0%, see **Fig. 5C** for flowchart). These 136 participants, of which 54 (39.7%) were female and 82 (60.3%) were male, had a median age of 55 (IQR: 41-66) years (see **Table 2** for population characteristics).

**Table 2.** Population characteristics of serologically tested individuals participating in the online health survey.

|  |  |
| --- | --- |
| Individuals, number | 136 |
| Median age (IQR), years | 55 (41 to 66) |
| Sex, female | 54 (39.7%) |
| Sex, male | 82 (60.3%) |

71 individuals (52.2%) had a TRABI-based posterior probability ≥ 0.5 and were considered seropositive, 65 (47.8%) had a posterior < 0.5 and were considered seronegative. Within the seronegative population, 98.4% reported no infection prior to blood sampling, while 53.5% of the seropositive individuals reported a known prior infection (**Table S3**). At the time of blood sampling, the agreement between seropositivity and knowledge of infection was moderate (Cohen’s Kappa 0.51, percent agreement 74.8%). Over the full timeframe since the start of the pandemic, 77.6% (52/67) of seropositive individuals and 44.4% (28/65) of seronegative individuals reported an infection up to April/May 2022. To explore the potential effects of CoV2 infection on participants’ post COVID-19 health status, we focused on these 80 individuals reporting an infection, using the 56 individuals without known infection as a comparison.

Amongst those with known CoV2 infection up to April/May 2022, 81.0% reported one or multiple symptoms at the time of infection, while 19.0% reported asymptomatic infection; a result that is consistent with findings by others (Ma *et al*., 2021; Sah *et al*., 2021). Cough, fatigue, and fever were the three most frequent symptoms that were reported during acute infection (**Fig. 5D**). We next assessed the time between the first reported infection and survey completion. The median time since first infection dated back 525 (IQR:57-571) days and the time frame included three pandemic peaks (**Fig. 5E**): in Spring/Summer 2020 (first WT CoV2 variant wave), in Fall/Winter 2020/2021 (second WT CoV2 variant wave), and in Winter/Spring 2021/2022 (omicron CoV2 variant wave).

The proportion of hospitalized individuals decreased with time (41.7% in Spring/Summer 2020, 23.5% in Fall/Winter 2020/2021 and 2.9% in Winter/Spring 2021/2022), with diagnosed pneumonia being more frequent in Spring/Summer 2020 (25.0%) than in Fall/Winter 2020/2021 (15.2%) and Winter/Spring 2021/2022 (3.0%, see **Table S4**).

In terms of recovery, 56.9% of the study participants with known infection by April/May 2022 stated to have fully recovered to their normal health status (45.5% in infected during first wave in Spring/Summer 2020, 61.3% in infected during Fall/Winter 2020/2021, 56.7% in infected during Spring 2022). Overall 9.8% reported that they were still experiencing at least some of the initial symptoms at the time of survey completion. 90.2% stated that symptoms lasted up to 3 months, with no study participant experiencing symptoms lasting between 3 and 6 months. Among those infected with WT CoV2, 11.4% reported that they were still experiencing symptoms more than 12 months after infection. The proportion with ongoing symptoms was comparable between infection waves, albeit slightly lower for the omicron wave (10.0% in Spring/Summer 2020, 13.0% in Fall/Winter 2020/2021 and 7.1% in Winter/Spring 2021/2022). Three individuals (8.3% of those with known infection during the first two waves) reported to have been diagnosed with post COVID-19 condition (long COVID-19).

The prevalence of symptoms within the past seven days (before completing the survey) among the previously infected group was highest for fatigue, sleeping problems, reduced performance, cough and concentration (**Fig. 5F**). Meanwhile, when comparing symptom prevalence among previously infected with those that had never experienced an infection, cough, gastrointestinal symptoms, skin problems, nervousness, myalgia, arthralgia and depression were reported more frequently by participants, among others (**Fig. 5G**, logistic regression, adjusted for age and sex). However, these differences did not reach statistical significance, with the exception of cough (odds ratio = 10.7, p-value = 0.026, adjusted for age and sex). A higher number of participants would likely clarify on some of the trends observed here.

We next asked the patients to report on new medical diagnoses that they have obtained after 2020. Here, we aimed to find out whether the prevalence of disease classes was fundamentally different in patients after infection with CoV2, while using the non-infected group as control. The most commonly medically diagnosed conditions of those with infection were related to skin, lung, thyroid, kidney, and immune system (**Fig. 5H**), while none of the comparisons with the non-infected group reached statistical significance (logistic regression, adjusted for age and sex). Of note, those who got infected during the first wave displayed a particularly high frequency of neurological diagnoses, and a comparatively low proportion of participants with new medical diagnoses was observed in those infected during the Winter/Spring 2021/2022 wave.

Then, we assessed the participants’ health status using the EuroQol 5-dimension 5-level instrument (EQ-5D-5L) and the EuroQol visual analogue scale (EQ VAS), where increased EQ-5D-5L and EQ VAS scores correspond to increased/better health. Overall, there was no statistically significant difference in EQ-5D-5L and EQ VAS scores between individuals reporting a known infection (mean EQ-5D-5L: 0.87, standard deviation (SD): 0.19; mean EQ VAS: 75.00, SD: 15.83) than those not infected (mean EQ-5D-5L: 0.81, SD: 0.17, p-value=0.13; mean EQ VAS: 70.30, SD: 20.88, p-value=0.15; logistic regression, adjusted for age and sex; see **Table S5**).

Lastly, we repeated these analyses to compare the longer-term health impacts between individuals with symptoms during acute infection (n=64) and individuals with asymptomatic infection (n=30). Both symptoms experienced during the last seven days (**Fig. 5I**) as well as new medical diagnoses (**Fig. 5J**) did not display statistically significant differences between the two groups. Similarly, EQ-5D-5L and EQ VAS scores between symptomatic (mean EQ-5D-5L: 0.87, SD: 0.19; mean EQ VAS: 77.54, SD: 11.73) and asymptomatic individuals (mean EQ-5D-5L: 0.86, SD: 0.17; mean EQ VAS: 69.00, SD: 21.41) did not differ significantly (p-value=0.879 for EQ-5D-5L and p-value=0.02 for EQ VAS; logistic regression, adjusted for age and sex). Due to the limited sample size, the findings regarding symptoms, new medical diagnoses and longer-term health impairment need to be interpreted with caution. We found no evidence for a difference in longer-term health outcomes between individuals with symptomatic and asymptomatic acute infection. These results suggest that post COVID-19 condition, with symptoms lasting longer than twelve months, occurs in approximately 10%.

### Prevalence of anti-CoV2 antibodies in prepandemic samples

5’475 prepandemic plasma samples (4’379 USZ patients and 1’096 healthy blood donors) were examined for the presence of cross-reactive antibodies against S, RBD and NC of CoV2. Several individuals had a strong antibody response against a single antigen and an absence of binding to other antigens, reflected in a low posterior probability but high –log(EC_50_) value. We then directly compared prepandemic and copandemic samples in the USZ cohort on the basis of single antigens and their respective posterior probabilities. When focusing on samples with high values for single assays, we observed an enrichment of high posterior probabilities in pandemic but not in the prepandemic group (**Fig. 6A**). Among samples with individual –log(EC_50_) values above 2 in May and June 2020, 76% (S), 80% (RBD), and 22% (NC) had a posterior probability > 0.5. In the prepandemic samples, maximally 1 sample with an individual assay level above 2 had a posterior probability above 0.5. This enrichment is suggestive of a substantial performance improvement when using the combined metric in the USZ cohort.

**Fig. 6.**
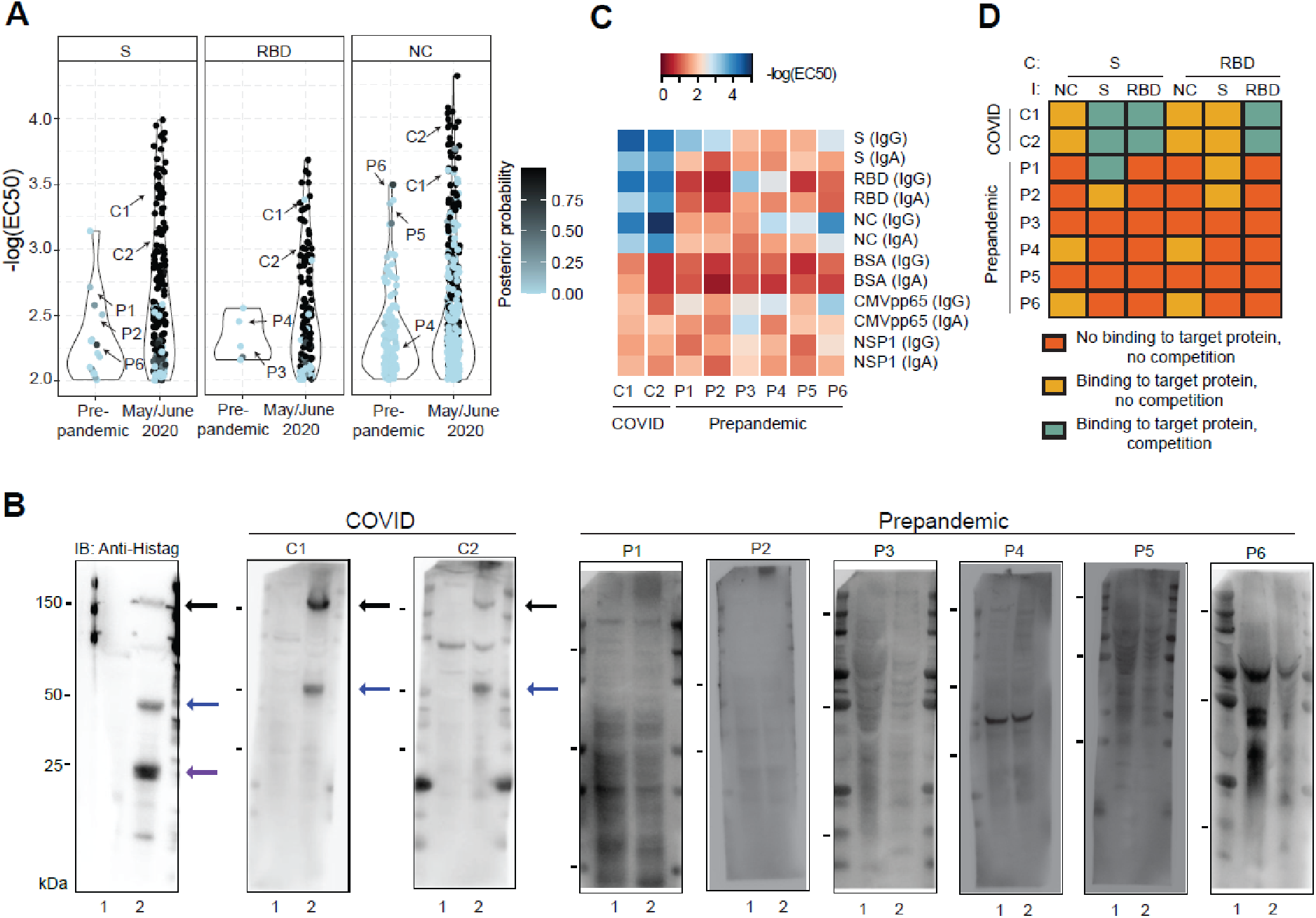
Characterization of prepandemic samples. **A**. Posterior probability were calculated assuming a Gaussian distribution and visualized for individual antigens (S, RBD and NC) for prepandemic samples vs. copandemic USZ samples drawn in May and June 2020. Prepandemic samples exhibited a low posterior probability as they typically reacted against single antigens, leading to low rankings in a composite metric. For further testing, comparative samples were chosen from the prepandemic era and from May and June 2020. Arrows point to samples of individuals used in (B), (C), (D). P1-6: prepandemic 1-6; C1-2: COVID1-2. **B**. Western Blot analysis of two samples from May/June 2020 (“COVID 1” or C1 and “COVID 2” or C2) and several prepandemic samples (P1-6). Anti-his-tag antibody was included as a positive control. Lane 1 = non-transfected Expi293F cell lysate; Lane 2 = Expi293F cell lysates expressing his-tagged S, NC and RBD proteins. Black arrows: S; blue arrows: NC; purple arrow: RBD. **C.** ELISA assays on the same samples as in B, using CoV2 S, NC, RBD and NSP1 as well as control proteins (BSA, CMV pp65), shown in the form of a heatmap where the -log(EC_50_) of the sample dilution is depicted. **D**. Competition assays were carried out in the same samples as in B and C. Competition (C) was performed with S (0.04-88 nM) or RBD (0.7-1350 nM) and plates were immobilized (I) with S, RBD, or NC. Data from duplicates is depicted using the following qualitative categories: No binding to target protein, no competition (orange). Binding to target protein, no competition (yellow). Binding to target protein, competition (turquoise). Soluble antigens suppressed the ELISA signal in the COVID samples but not in the prepandemic sample (except for P1 where soluble S competed the immobilized S), showing that the antibodies present in the latter had lower affinities for CoV2 targets.

We then compared the immunochemical properties of six prepandemic samples with high binding to S, RBD or NC to two samples of confirmed COVID-19 (COVID 1 and 2, see annotation in **Fig. 6A**). The COVID-19 samples, but not the prepandemic samples, recognized in Western blots the S and NC antigens of CoV2 expressed by Expi293F (**Fig. 6B**). Additional ELISAs performed on the same samples confirmed the initial findings (**Fig. 6C**) including intact binding to the RBD. The discrepancy between ELISA and Western Blot suggests that the RBD is a highly conformational epitope lost upon boiling and SDS denaturation.

To further probe the specificity of the findings, we also carried out competitive ELISAs on prepandemic and COVID patients. First, we determined plasma concentrations close to the EC_50_. Then we pre-incubated appropriately diluted samples with various concentrations of S and RBD (0.04-88 and 0.7-1350 nM, respectively). Samples were then transferred onto ELISA plates coated with S, RBD, and NC. The concentration-dependent displacement of the measured optical density was then interpreted and categorized into three distinct classes: (1) No binding to the target protein, no competition. (2) Binding to the target protein, no competition. (3) Binding to the target protein, competition (**Fig. 6D**). We found that both soluble S and the RBD caused a concentration-dependent depletion of the RBD in COVID samples. The S signal could not be depleted with RBD, indicating the presence of epitopes other than the RBD. One prepandemic sample (#1) displayed competition of the S signal with soluble S but not with soluble RBD. Other prepandemic samples did not show competition at all, suggesting that their reactivity was due to high concentrations of low-affinity antibodies cross-reacting with CoV2 S.

### Identification of seropositives in healthy donors and clonality of anti-S immune response

TRABI enabled the identification of 189 CoV2 seropositive blood donors that underwent regular blood donation at the blood donation service of Zurich (**Fig. 2B**, **C**) despite clear serological indications of past infection and antibody titers in the same range as those of PCR-confirmed convalescent individuals (**Fig. 7A**). We assessed IgG and IgA antibodies to S, RBD, and NC as well as responses to multiple control antigens, in 4 healthy blood donors and 4 convalescent individuals recruited to the BDS. We observed binding of IgG antibodies in blood donors and convalescent individuals against S, RBD, and NC, with usually lower IgA titers. No binding against the CoV2 non-structural-protein 1 (NSP1), or against bovine serum albumin (BSA) was observed.

**Fig. 7.**
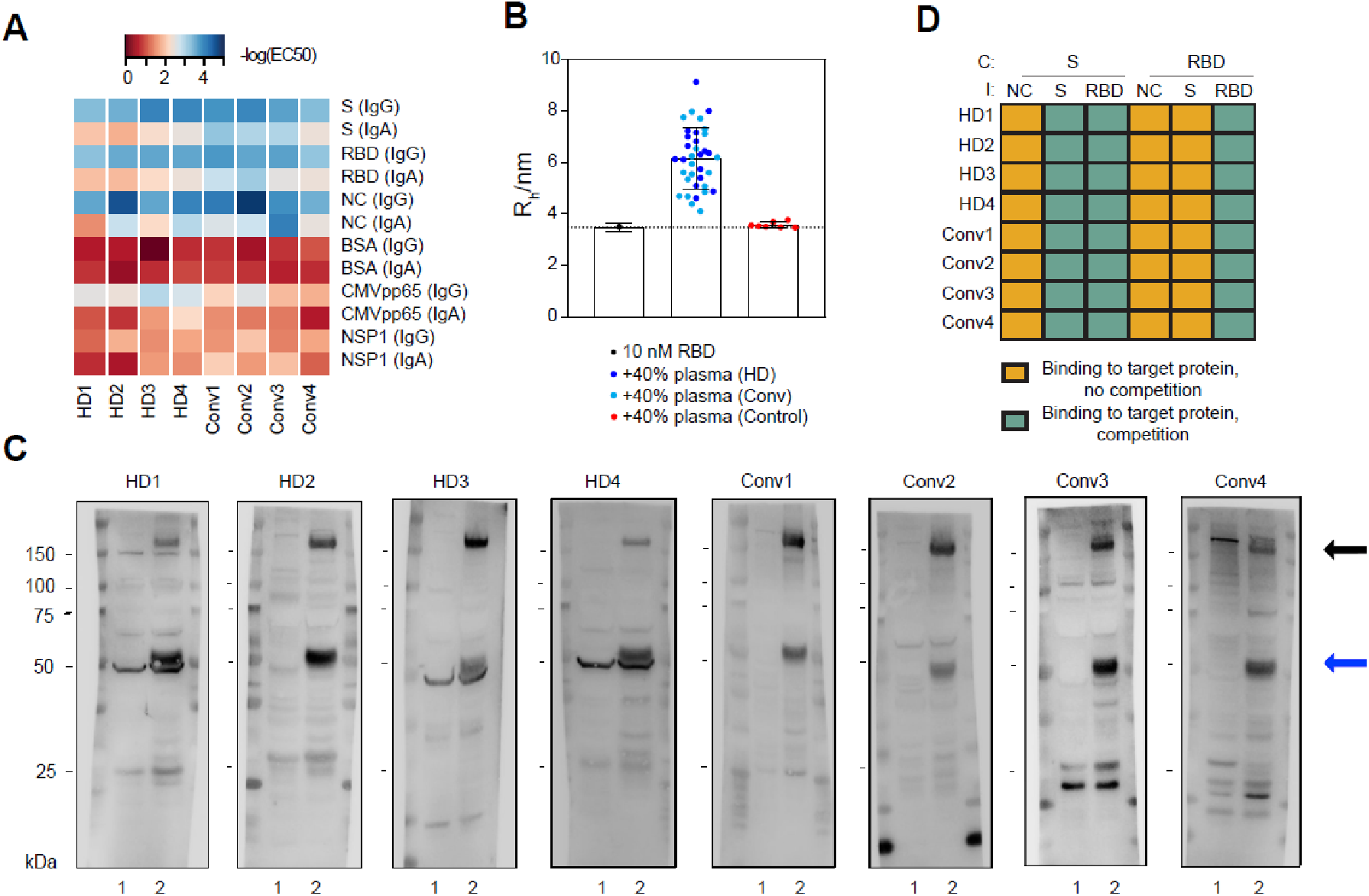
Assay validation in solution and clonality of anti-S immune response. **A**. ELISA assays of healthy blood donors vs. convalescent individuals depicted as heatmap. The -log(EC_50_) depicts the sample dilution at which half-maximum binding occurs. S, RBD, and NC are strongly bound by both healthy donors (HDs) as well as convalescent (Conv) individuals. **B**. Microfluidic-based assessment of binding between an Alexa 647-labelled RBD antigen and antibodies in solution. No change in diffusion coefficient or the associated hydrodynamic radius was observed in control samples, while all ELISA-positive samples from convalescent and healthy donors indicated a clear binding of antibodies to RBD, confirming the ELISA-based results. **C**. Western Blot analysis of the same individuals tested in (A). Lane 1 = non-transfected Expi293F cell lysate; Lane 2 = Expi293F cell lysates expressing his-tagged S, NC and RBD proteins. Black arrows: S. Blue arrows: NC. **D**. Competitive ELISA using RBD or S for soluble competition with antibodies in plasma from the same individuals as in (A) and (C). Data is depicted using the following qualitative categories: Binding to target protein, no competition (yellow). Binding to target protein, competition (turquoise). Competition (C) with S or RBD did not change the signal upon immobilization (I) with NC, while competition with S resulted in a decrease in signal upon immobilization with S as well as with RBD. Conversely, competition with RBD only competed signal when immobilized with RBD, not with S, indicating the presence of antibodies against S domains other than RBD.

To further validate the seropositivity in healthy blood donors, we employed an orthogonal methodology which allows antibody/antigen interactions to be probed in solution, without any immobilization of antigens to a surface (Schneider *et al*., 2022). Samples of CoV2 convalescent individuals, healthy donors and controls were preincubated with fluorescently conjugated RBD protein. We then monitored the increase in the effective molecular weight of an Alexa647-labelled RBD construct in solution upon complex formation with an antibody present in the patient sample. This was achieved by measuring the associated decrease in its molecular diffusion coefficient upon binding using a microfluidic platform. While no change in diffusion coefficient or the associated hydrodynamic radius was observed in control samples, all ELISA-positive samples from convalescent and healthy donors indicated a clear binding of antibodies to RBD (**Fig. 7B**). We confirmed these findings by using the samples of several healthy blood donors and convalescent individuals as primary antibodies in Western Blot and detected bands for both S and the NC in the Expi293 cells overexpressing the viral proteins but not in the Expi293 control lysate (**Fig. 7C**).

To obtain a rough estimate of the clonality and epitope specificity of the immune response raised against the S protein, we conducted an ELISA-based soluble antigen competition. Competition with the RBD lead to a decrease in ELISA signal for RBD but not for S or NC in both convalescent individuals and healthy blood donors (**Fig. 7D**). Conversely, competition with S decreased the signal for both S and the RBD, suggesting the presence of antibodies targeting multiple S epitopes, including RBD. Therefore, the immune response against S was polyclonal and involved multiple viral epitopes.

## Discussion

Using a high-throughput CoV2 serology pipeline, we draw a detailed picture of the evolution of CoV2 seroprevalence in a large central-European metropolitan area. If antibody titers were stable after infection, the seroprevalence would reflect the entirety of the population infected since inception of the pandemic. However, anti-CoV2 titers were found to decay in multiple studies (Buss *et al*., 2020; Ibarrondo *et al*., 2020; Q. X. Long *et al*., 2020; Seow *et al*., 2020; Ward, Cooke, *et al*., 2020; Choe *et al*., 2021), with a half-life of approximately 106 (CI95% 89 to 132) days (Buss *et al*., 2020), and others suggesting an even shorter half-life of 26 to 60 days (Ibarrondo *et al*., 2020). This decrease in titers over time was confirmed in neutralization assays, shown in various studies (Q. X. Long *et al*., 2020; Seow *et al*., 2020; Choe *et al*., 2021). Indeed, between April and July 2020 the prevalence of seropositivity fell by ≈60% in our cohorts, which confirms the waning of humoral immunity at the population level. Using an extended SEIR model, we estimated that the population-wide half-life of seropositivity is 75 (CI95% 55-103) days (unadjusted seroprevalence data) or 88 (CI95%: 61-128) days (after post-stratification for age and sex).

If our sampling methodology suffers from systematic errors, the cohorts sampled here may not be representative of the population studied. In order to minimize such issues, we surveyed two non-overlapping cohorts: hospital in- and outpatients and healthy blood donors. Neither cohort can be assumed to represent a representative random sample of the population. However, poststratification by age and sex led to only minor changes in seroestimates, indicating that our cohorts are largely representative of the population of the canton of Zurich. However, we have not investigated the extent of CoV2 spread in children in the canton of Zurich, which was recently done by others (Ulyte, 2020).

The dynamics of the seroepidemiology confirms that the outbreak followed three distinct phases. The cumulative incidence rose during the first wave in spring 2020, with 2.3% (CrI95%: 2.0%- 2.8%) having contracted CoV2 by June 2020. There was a modest increase over the summer months, followed by a rapid rise in late 2020. We estimate that 10.3-14.6% had undergone an infection with CoV2 by mid-December 2020.

Thereby, we could delineate the precise serological status in the population of the canton of Zurich in a continuous manner, rather than on single points in time. These estimates of CoV2 antibodies were performed on a highly sensitive immunoassay (TRABI) that combines antibody measurements against three CoV2 proteins in a QDA-based compound metric, a system developed in house. In view of the critique levelled at past serological studies (Bendavid *et al*., 2020; Streeck *et al*., 2020), we have gone to great lengths to assess and validate our technology, using several orthogonal techniques. A recent publication (Ng *et al*., 2020) has shown pre-existing anti-CoV2 antibodies in unexposed humans. Antibody sizing (Arosio *et al*., 2016; Schneider *et al*., 2022) and immunoblots, however, point to fundamental differences between prepandemic seropositivity and the immune responses of CoV2-infected individuals. While the latter consistently showed high-affinity responses that were clearly visible in Western blotting, the few seropositive prepandemic sera were unanimously negative in Western blotting, and equilibrium displacement ELISA of one prepandemic plasma sample suggested a much lower affinity despite similar antibody EC_50_ titers. We conclude that any immune response in uninfected individuals, whether it represents cross-reactivity with common-cold coronaviruses or something else, is of inferior quality and may less likely be protective. A blinded comparison with commercial test kits showed that our approach was suitable for large-scale epidemiologic studies and that the compound metrics did indeed lead to a power gain, as shown by the enrichment of samples with high posterior probabilities in excess of the single assays during the epidemic.

The comparably low seroprevalence of CoV2 in the canton of Zurich, in particular during the first wave, is compatible with other more affected regions, based on the reported IFR, in Switzerland (Perez-Saez *et al*., 2020) and in European areas with similar medical infrastructure (Hauser *et al*., 2020). While some large-scale serological surveys performed throughout the globe revealed CoV2 spread slightly exceeding the values we observed in Zurich (Le Vu *et al*., 2020; Pollán *et al*., 2020; Ward, Atchison, *et al*., 2020), other studies identified regions with seroprevalence surpassing 50%, e.g. in some areas in the Amazonas state in Brazil (Buss *et al*., 2020) or in slums in Mumbai, India (Malani *et al*., 2020). Yet, since antibody waning has been reported in multiple instances (Buss *et al*., 2020; Ibarrondo *et al*., 2020; Q. X. Long *et al*., 2020; Seow *et al*., 2020; Ward, Cooke, *et al*., 2020; Choe *et al*., 2021), discrete seroestimates may reflect snapshots of the immunity status of a population at a certain time, rather than the true cumulative case incidence. Conversely, we have accounted for antibody waning, using a model fit developed by data obtained through continuous CoV2 seromonitoring. Thereby, we were able to derive the cumulative incidence rate for both the first and the second wave of the epidemic in the canton of Zurich and have shown that the nation-wide antigen testing underestimates the true number of CoV2 infections by approximately factor 3, similar to what was found in France (Pullano *et al*., 2021).

By now, vaccination campaigns in the canton of Zurich, throughout Switzerland, and in multiple places across the globe have rapidly advanced, reaching a stage where novel booster candidates (e.g. mRNA-1273.211, a bivalent booster vaccine candidate), with expected superior activity against many known variants of concern, become available. Yet, the continuous monitoring of the antibody response will remain a crucial component to epidemiologically assess the extent of immunity within our population over time (Altmann and Boyton, 2020; Quast and Tarlinton, 2021), in children as well as in adults. Our TRABI assay may be particularly meaningful since we can distinguish between natural infections (eliciting an antibody response also against the NC protein) and vaccination-induced immunity (targeting the S protein). Our cohort of hospital patients will be further surveyed for the surge of unexpected clinically relevant sequalae that may be associated with an infection of CoV2. Initial analyses performed on our dataset did not reveal clusters of disease entities associated with CoV2 infection, compared with patients with no history of CoV2 seropositivity. Along these lines, our data does not indicate an increased prevalence of Parkinson’s Disease upon CoV2 infection, an association suggested by recent case reports (Cohen *et al*., 2020; Faber *et al*., 2020; Méndez-Guerrero *et al*., 2020). Interestingly, male patients were overrepresented in the cohort with severe disease requiring hospitalization although infections seems to be roughly equally distributed between female and male. As a clear limitation of our approaches, maladies that do not require treatment at a university hospital center may be altogether missed since the patients may be referred to a practitioner outside the university setting, whereby the occurrence of disease would not be entered in the hospital database system. Moreover, pseudonymized records of patient data, used for the protection of sensitive information from patients, do not allow to gain access to detailed non-parameterized files and the presence of a diagnosis may be missed.

Meanwhile, we were able to provide additional depth regarding the post COVID-19 health status of patients whom we had identified as seropositive using the TRABI assay or who self-reported an infection with CoV2 up to April/May 2022 through a standardized online health survey. We found that 11.4% of those reporting an infection in the first two pandemic waves (Spring/Summer 2020 and Fall/Winter 2020/2021) still complained about ongoing symptoms after >12 months after infection, and 8.3% had received a diagnosis of post COVID-19 condition (‘long COVID’). While numbers in the literature cover a wide range of about 14-75% potentially affected by post COVID- 19 condition up to one year after diagnosis (Huang *et al*., 2021; Menges, Ballouz, *et al*., 2021; Sigfrid *et al*., 2021; Bull-Otterson *et al*., 2022; Heesakkers *et al*., 2022; Nehme *et al*., 2022), our findings are comparable to those of other population-based studies (Menges, Ballouz, *et al*., 2021; Ballouz *et al*., 2022). Online health survey participants with known infection reported several symptoms and new medical diagnoses more frequently than those without infection, but differences were not statistically significant and no differences in health status (EQ-5D-5L and EQ VAS) were observed between these groups. Similarly, no significant differences in long-term out-comes between individuals with symptomatic and asymptomatic infection were identified, suggesting that the occurrence of post COVID-19 may be independent of symptoms during acute infection. Yet, these analyses are limited by the participation rate resulting in a relatively small sample size. Certain consequences of CoV2 infection – potentially CoV2 clade dependent (Emmenegger *et al*., 2022) – may take more time to manifest and large numbers of patients may need to be assessed to perform solid statistical analyses due to the heterogeneous clinical picture (Gavriatopoulou *et al*., 2020; Kordzadeh-Kermani, Khalili and Karimzadeh, 2020; Pezzini and Padovani, 2020; Emmenegger, Saseendran Kumar, *et al*., 2021) and the phenotypic heterogeneity of post-acute COVID-19 sequalae (Rando *et al*., 2021). Furthermore, it cannot be excluded that selection effects or potential residual confounding may have influenced the findings of the survey. However, our key findings – emerging from comparisons with a much needed control group (‘never infected’) often omitted in observational studies (Aschman *et al*., 2022) – are consistent with the literature and underpin that longer-term symptoms and complications post COVID- 19 are an important concern for patient care and public health.

Ultimately, as much of a catastrophe as CoV2 has been, we are not immune to future epidemic outbreaks of other viral diseases potentially far worse. Yet, a multidimensional, comprehensive understanding of CoV2 may provide crucial epidemiological tools to prevent an epidemic at an early stage, to save lives and increase life quality throughout the world.

## Materials and Methods

### Ethics

All experiments and analyses involving samples from human donors were conducted with the approval of the ethics committee of the canton Zürich, i.e. Kantonale Ethikkommission Zürich (KEK-ZH-Nr. 2015-0561, BASEC-Nr. 2018-01042, and BASEC-Nr. 2020-01731), in accordance with the provisions of the Declaration of Helsinki and the Good Clinical Practice guidelines of the International Conference on Harmonisation. All human donors and patients included in this study provided a written general or informed consent.

### Study design and sampling

The seroepidemiological survey of CoV2 infection in the greater area of Zurich is a population-based study to investigate the temporal evolution of seropositivity for CoV2 in two independent cohorts. We made use of surplus plasma samples from inpatients and outpatients admitted to the University Hospital of Zurich (USZ) collected daily (Monday-Friday) and used for population-wide interrogations of the antibody repertoire (Senatore *et al*., 2020). For the CoV2 seroprevalence study, we included 4’379 samples prior to December 2019 (prepandemic samples) and 51’435 samples from December 2019 to December 2020 (copandemic samples). The criteria for our study to include a sample into the analysis were: (1) The patients’ blood was sent to the Institute of Clinical Chemistry (at USZ), (2) there was enough residual heparin plasma (150 µl) for the automated generation of a research aliquot, (3) no aliquot from the same patient was already provided within the same month, (4) additional information (age, sex, clinical ward to which patient was admitted) was available. Point (3) led to the exclusion of 415 samples and point (4) to the exclusion of 30 samples for the calculation of the seroestimates. While not being representative for the population of the canton of Zurich sensu stricto, we have selected this patient cohort due to the depth of available medical data that will allow to trace long-term effects of CoV2 infections from a clinical stance. At the same time, many of the hospital patients are among the most susceptible within a population and are thus in need of substantial monitoring.

Similar to others (Buss *et al*., 2020; Filho *et al*., 2020; Slot *et al*., 2020; Uyoga *et al*., 2020; Xu *et al*., 2020), we have investigated CoV2 IgG seroprevalence of a healthy adult population, complementing the hospital patients, in blood donors of the Blood Donation Service of the Canton of Zurich. Overall, 16’291 samples (thereof 1’096 prior to December, 2019) from blood donors who consented to further use of their samples for research were randomly selected every month (on average: 1’170 samples/month from December 2019 to December 2020) and sent from the blood donation service to Neuropathology. The criteria to be admitted for blood donation are in line with international standards of blood donation services, see (Blutspendedienst, 2021a). Blood donors with a confirmed CoV2 infection are excluded from donating blood for four weeks, following the full remission of symptoms. Blood donors have to be at least 18 years of age, weigh at least 50 kg, and feel healthy. In order to be included for blood donation, donors have not undergone a substantial surgery or pregnancy/birth in the past 12 months, have not been subjected to dental treatments in the past 72 hours, and have not received foreign blood since 01.01.1980. Moreover, the inclusion mandates that blood donors have not been to an area at risk of malaria or another region with a high prevalence of infectious diseases. Blood donors are only admitted if they have not been tattooed or acquired a permanent make-up in the past four months. A positive test for HIV, syphilis, hepatitis C or B leads to a definite exclusion. Additionally, blood donors are excluded if they have had new sexual partners within the last four months and if they display sexual risk behavior. Lastly, donors have not been to the England, Wales, Scotland, Northern Ireland, Isle of Man, Channel Islands, Gibraltar or to the Falkland Islands for more than six months between 1980 and 1986. Blood donors over age 65, until maximally age 75, can continue donating blood if they have donated blood earlier (the last, complication-free donation has to date back no longer than two years) and the health survey does not indicate any particular health risk. The detailed inclusion and exclusion criteria are enumerated here (Blutspendedienst, 2021b). Specimens were denoted according to the following conventions: *prepandemic samples*: samples collected before December 2019; *COVID samples*: samples from patients with clinically and/or virologically confirmed CoV2 infection; *copandemic samples*: any samples collected in December 2019 or thereafter.

### High-throughput serological screening

In order to test the samples for the presence of IgG antibodies directed against CoV2 antigens, high-binding 1536-well plates (Perkin Elmer, SpectraPlate 1536 HB) were coated with 1 µg/mL S or RBD or NC in PBS at 37 °C for 1 h, followed by 3 washes with PBS-T (using Biotek El406) and by blocking with 5% milk in PBS-T (using Biotek MultifloFX peristaltic pumps) for 1.5 h. Three µL plasma, diluted in 57 µL sample buffer (1% milk in PBS-T), were dispensed at various volumes (from 1200 nL down to 2.5 nL) into pre-coated 1536-well plates using contactless dispensing with an ECHO 555 Acoustic Dispenser (Labcyte/Beckman Coulter). Sample buffer was filled up to 3 uL total well volume using a Fritz Gyger AG Certus Flex dispenser. Thereby, dilution curves ranging from plasma dilutions 1:50 to 1:6000 were generated (eight dilution points per patient plasma sample). After the sample incubation for 2 h at RT, the wells were washed five times with wash buffer and the presence of IgGs directed against above-defined CoV2 antigens was detected using an HRP-linked anti-human IgG antibody (Peroxidase AffiniPure Goat Anti-Human IgG, Fcγ Fragment Specific, Jackson, 109-035-098, at 1:4000 dilution in sample buffer). The incubation of the secondary antibody for one hour at RT was followed by three washes with PBS-T, the addition of TMB, an incubation of three minutes at RT, and the addition of 0.5 M H_2_SO_4_ (both steps with Biotek MultifloFX syringe technology). The final well volume for each step was 3 µL. The plates were centrifuged after all dispensing steps, except for the addition of TMB. The absorbance at 450 nm was measured in a plate reader (Perkin Elmer, EnVision) and the inflection points of the sigmoidal binding curves were determined using the custom designed fitting algorithm described below. The secondary antibodies we have used were tested and validated previously (Emmenegger *et al*., 2022) and replicability as well as influence of different sample types (e.g. serum and heparin plasma) on the TRABI have already been reported (Emmenegger, Saseendran Kumar, *et al*., 2021; Emmenegger *et al*., 2022).

### Counter screening using commercial and custom-designed platforms

We used the following commercial tests for the detection of anti-CoV2 antibodies in 55 plasma samples of 27 patients who were diagnosed by RT-PCR to be infected by CoV2 as well as 83- 90 plasma samples which were collected before December 2019 and, hence, before the start of the COVID-19 pandemics: The double-antigen sandwich electro-chemiluminescence immunoassay from Roche diagnostics (Rotkreuz, Switzerland) was performed with the E801 of the COBAS8000® system (Roche diagnostics, Rotkreuz, Switzerland). The test detects any antibody against the nucleocapsid antigen. The fully automated LIAISON® CoV2 chemiluminescence immunoassay from DiaSorin (Saluggia, Italy) detects IgG against the S1/S2 antigens. The CoV2 chemiluminescent microparticle immunoassay from Abbott (Abbott Park, IL, USA) detects IgG against the nucleocapsid antigen and was performed on an Architect™ analyser. Two ELISAs from EUROIMMUN (Lübeck, Germany) detect IgA or IgG against the S1 antigen and were performed by the use of a DSX™ Automated ELISA System (DYNEX Technologies (Chantilly, VA, USA). The high-throughput serology assay in Oxford (under development) was carried out in the Target Discovery Institute, University of Oxford. High-binding 384-well plates (Perkin Elmer, SpectraPlate) were coated with 20 µL of 2.5 µg/mL S o/n at 4°C, followed by 3 washes with PBS-T and by blocking with 5% milk in PBS-T for 2 h. Blocking buffer was removed and 20 µL of 1:25 sera diluted in sample buffer (1% milk in PBS-T) was dispensed into S-coated wells then incubated for 2 h at RT. The wells were washed five times with wash buffer and the presence of IgGs directed against S was detected using an HRP-linked anti-human IgG antibody (Peroxidase AffiniPure Goat Anti-Human IgG, Fcγ Fragment Specific, Jackson, 109-035-098) at 1:50,000 dilution in 20 µL sample buffer. The incubation of the secondary antibody for one hour at RT was followed by three washes with PBS-T and the addition of QuantaRed™ Enhanced Chemifluorescent HRP Substrate Kit (Thermo Scientific, Waltham Massachusetts, USA) then incubated for four minutes at RT before the addition of the stop solution. The fluorescence at excitation/emission maxima of ∼570/585nm was measured in a fluorescent plate reader (Perkin Elmer, EnVision).

### Analysis of data derived from high-throughput serological screen

#### Data fitting

Eight-dilution points equally spaced on a logarithmic scale are fitted with an equation derived from a simple binding equilibrium. The inflection point (-log_10_EC_50_) is extracted from the fit. Baseline and plateau values are fixed by the respective positive and negative controls in a plate-wise fashion and the signal is fitted following these equations:

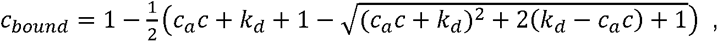

where c_bound_, c_a_ and c are concentration of the antigen-antibody, antigen, and blood concentration respectively.

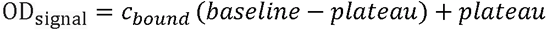

#### Data preprocessing

All samples that yielded an –log_10_EC_50_ of below –3 on any antigen were labelled as non-fittable and non-detectable. Their dilution curves cannot be differentiated from baseline and therefore only an upper bound for the –log_10_EC_50_ can be determined. These samples were therefore excluded from data fitting but were of course included in ROC analysis and prevalence estimation.

#### QDA, LDA, and Prevalence estimation

Assume that we have data for *m* samples with known serostatus and antibody measurements, that is, we have (*X*_*i*_, *Y*_*i*_), *i* = 1, . ., *m*, where *X*_*i*_ is the vector of size *p* (in our case our antigen measurements) and *Y*_*i*_ is a Boolean variable defining group membership (in our case, whether the individual is seropositive or not). The QDA model assumes multivariate normal distributed *X*_*i*_ given *Y*_*i*_:

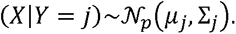

Further, the model assumes that the prior, that is, distribution of *Y*_*i*_, is known s. t. *P*[*Y* = *j*] = *π*_*j*_ The quadratic discriminant classifier simply assigns each sample to the group which has the larger posterior *P*[*Y*|*X*], which is proportional to the joint probability *P*[*Y*, *X*].

Therefore, we assign sample *i* to group 1 if

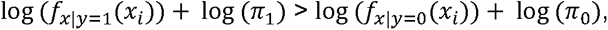

And to group 0 otherwise. To set the prior, one option is to take just the mean of the group sizes. However, this is not an ideal option in our case, where we have an additional *n* samples with unknown serostatus to classify: The prevalence in the *m* samples with known serostatus might deviate substantially from the prevalence in population with unknown serostatus. We therefore estimate *π*_1_directly from the data of unknown serostatus using a simple expectation maximization scheme. Proceeding in an iterative fashion, from a given estimate *π*_1_^*k*^, we define the posterior (E step):

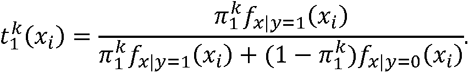

Then, we update our estimate of *π*_1_ (M step):

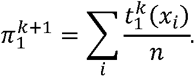

After convergence, this yields our estimate of the positive serostatus prevalence in the samples. Note that the sample ordering according to this classifier is independent of the prior and therefore has no impact on an analysis via ROC curves. Further, note that evaluating QDA via ROC analysis, an out of sample scheme should be employed to avoid biased estimates of performance; we chose 10-fold cross-validation throughout. Lastly, note that the strategy does not critically depend on the normality assumption but just requires an estimate for the density functions, *f*_*x*|*y*=*j*_(*x*_*i*_). Even nonparametric estimates could be an option.

For the LDA approach, we first collapse the antigen measurements per samples according to the linear discriminant classifier:

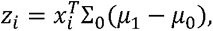

Where Σ_0_ is the covariance estimated from the known negatives only and *μ*_1_, *μ*_0_ are the means of the known positives and negatives respectively. The above algorithm is then applied on the resulting one dimensional variable *z*_*i*_. 95% confidence intervals were derived by bootstrap drawing 1000 bootstrap samples, where the number of samples drawn from each annotation group (known positives, known negatives and unannotated) was kept constant.

### Epidemiological modelling

#### Post-stratification of age and sex with distributional information of population of canton of Zurich

We adjusted the estimates for differences in age and sex between the population of the canton of Zurich and the samples of individuals hospitalized at UZH or blood donors using inverse probability weighting. The data for the population of the canton of Zurich were obtained from the statistical services of the canton of Zurich.

#### Antibody waning and cumulative incidence

Without presuming the effects of antibody waning on immunity to CoV2 re-infection, it is important to account for it when estimating the infection attack rate or cumulative incidence from seroprevalence data. To this aim, we propose an extension to the classic SEIR model where the R compartment (R for removed) in the classical formulation is split in 3: (1) Compartment R represents the subgroup of population that is removed from infectiousness and did not seroconvert yet. (2) Compartment A (for antibody) represents the sub- group of population that is removed from infectiousness and did seroconvert. (3) Compartment W (for waning) represents the subgroup of population that is removed from infectiousness and whose antibodies waned. We select a rate of 1/14 days for seroconversion (14 days on average from R to A) and estimate the rate of antibody waning from data. The model also assumes an average generation interval of 5.2 days (Ganyani *et al*., 2020), and an average time from disease onset to death of 20.2 days (Linton *et al*., 2020). We include a time-varying transmission factor by month, with smooth transitions handled by logistic switch functions. The model is fitted to seroprevalence data from USZ and BDS jointly, and to weekly mortality data from canton of Zurich with an infection fatality ratio fixed over time with a prior distribution set to 0.5% (95% central range: 0.2 to 1.0%) (Hauser *et al*., 2020). The model was fitted in a Bayesian framework using Stan (Carpenter *et al*., 2017). From the fitted model we can estimate the rate of antibody waning (or its half-life) as well as the infection attack rate/cumulative incidence corrected for antibody waning at any time point (1-S(t)).

#### Exploratory correlation analysis of CoV2 seropositivity with ICD-10 codes using Bayesian logistic regression

We explored associations between the posterior probability of a positive serology in individuals consulting at USZ and medical conditions as measured by the ICD-10 codes entered by the medical encoders for health insurance-related purposes. Whenever available, ICD-10 codes were extracted from our clinical data warehouse for all patients included in this study. Up to 100 different ICD-10 codes per case were annotated in a pivot table. We considered only the highest posterior probability for patients with multiple samples (some of which may be negative in the beginning and turn positive later on), and ICD-10 codes entered at any point. We limited the analysis to ICD-10 codes present in more than 0.1% of cases. The analysis was thus focused on 37’382 individuals and 199 variables, including age, sex and 197 ICD-10 codes. We used multiple logistic regression after logit-transforming the posterior probability. We placed ourselves in a Bayesian framework and conduct the analyses in the R package rstanarm (Goodrich *et al*., 2020). We started with standard regression, using uninformative priors on regression coefficients (Normal(0,10)). With this large number of covariates, the estimates were, as expected, very noisy and basically unusable. We thus used regularization techniques (Bayesian LASSO and regularized horseshoe priors, see (Gelman *et al*., 2013; Piironen and Vehtari, 2017)). We then showed the top five positive or negative associations between ICD-10 codes and posterior probability of CoV2 seropositivity (odds ratio with 95% credible interval).

#### Investigation of feature dissimilarity between seropositive and seronegative patients using linear and nonlinear dimensionality reduction mechanisms

The same dataset as described above (Bayesian logistic regression) was subjected to dimensionality reduction, with the following deviations: (1) Age was not included as a feature. (2) Seropositivity was defined as posterior probability ≥ 0.5. *PCA*. PCA was carried out using the default implementation in the R stats package (prcomp) and data was visualized using the factoextra package (https://cran.r-project.org/web/packages/factoextra/index.html). *UMAP*. The following UMAP configuration parameters from the umap package in R (https://CRAN.R-project.org/package=umap) were used, all of which are default, except for the metric where cosine was used instead of Euclidean due to the binary nature of the data (n_neighbors: 15, n_components: 2, metric: cosine, n_epochs: 200, input: data, init: spectral, min_dist: 0.1, set_op_mix_ratio: 1, local_connectivity: 1, bandwidth: 1, alpha: 1, gamma: 1, negative_sample_rate: 5, a: NA, b: NA, spread: 1, random_state: NA, transform_state: NA, knn: NA, knn_repeats: 1, verbose: FALSE, umap_learn_args: NA). UMAP data was plotted using ggplot2 in R.

#### Exploratory network analysis of ICD-10 codes, clinical departments, age, and sex for seropositive and seronegative patients

Topological networks have been constructed using the Cytoscape version 3.8.2 (https://cytoscape.org), to visualize the patient-ICD-10 code relationship on the network level (Jeong *et al*., 2017) and topological similarities between seropositive and seronegative USZ patients have been scored using the Mcode algorithm (Bader and Hogue, 2003). ICD-10 codes were depicted as purple rectangles, male patients as diamonds and female patients as circles. The serological status is encoded in red (seropositive) and blue (seronegative). A force directed layout was employed to represent the network.

#### Assessing potential complications of CoV2 infection in three patient groups using ICD-10 codes and free-text medical reports

Reports on complications of CoV2 infections beyond the classical COVID-19 pneumonia have accumulated over the past year. To investigate whether a CoV2 infection is associated with diseases that have not been linked to the virus so far, we first split our dataset into (1) seropositive COVID-19 patients hospitalized in the Infectious Diseases or Internal Medicine units (n=240, group I), (2) seropositive patients associated with other clinical wards (n=494, group II), and (3) randomly selected seronegative patients (n=635, group III). Group I likely reflects the cases hospitalized because of COVID-19, while group II is comprised of USZ patients that likely did not require hospitalization due to COVID-19 and some of the patients in this group may have been asymptomatic or paucisymptomatic. Our SQL databases containing ICD-10 codes and free-text medical reports were then queried individually for the three groups, using the following disease classes/conditions: 1) CoV2-related diseases. ICD-10 codes: J80, U69.0-!, J96%. Free text: ARDS, COVID-19-Pneumonie, respiratorische Insuffizienz, Dyspnoe, Lungenembolie. 2) Risk factors for severe disease/hospitalization (Jehi *et al*., 2020; Petrilli *et al*., 2020; Cummins *et al*., 2021). Free-text: Diabetes mellitus, Diabetes, Obesity, Herz-Kreislauf, Obesität, Hypertonie, COPD, Arrythmie, Arrythmia, chronische Nierenerkrankung, ischämische, Übergewicht, chronische Atemwegserkrankung, Bluthochdruck, Herzfehler, Herzversagen, chronic kidney disease. 3) Mixed neurological/neuropsychiatric (Almqvist *et al*., 2020). Free-text: Fatigue, Müdigkeit, Geschmack, Geruch, Verwirrung, Schwindel, Mood, Psychose, Enzephalitis, microbleed, Schlaganfall, Enzephalopathie, Delir, Epilepsie. 4) Extrapyramidal and movement disorders, therein Parkinson’s Disease. ICD-10: G20, G21, G22, G23, G24, G25, G26. Free-text: Parkinson, Dystonie, extrapyramidal, Chorea. 5) Inflammatory diseases of the central nervous system, therein encephalitis. ICD-10: G00, G01, G02, G03, G04, G05, G06, G07, G08, G09. Free-text: Enzephalitis, Enkephalitis, Enzephalomyelitis, Phlebitis, Meningitis, Myelopathie. 6) Demyelinating diseases of the central nervous system, therein multiple sclerosis. ICD-10: G35, G36, G37. Free-text: Multiple Sklerose, Demyelinisation, Demyelinisierung, Hirnsklerose. 7) Hypertensive diseases. ICD-10: I10, I11, I12, I13, I15. Free-text: essentielle Hypertonie, Bluthochdruck, Hypertensive Herzkrankheit, Hypertensive Nierenkrankheit. 8) Ischemic heart diseases.

ICD-10: I20, I21, I22, I23, I24, I25. Free-text: Angina pectoris, Myokardinfarkt, ischämische Herzkrankheit. 9) Pulmonary heart disease and diseases of pulmonary circulation. ICD-10: I26, I27, I28. Free-text: Lungenembolie, Lungeninfarkt, pulmonale Herzkrankheit, Thromboembolie. 10) Other forms of heart disease. ICD-10: I30, I31, I32, I33, I34, I35, I36, I37, I38, I39, I40, I41, I42, I43, I44, I45, I46, I47, I48, I49, I50, I51, I52. Free-text: Perikarditis, Perikarderguss, Endokarditis, Mitralklappenkrankheit, Pulmonalklappenkrankheit, Trikuspidalklappenkrankheiten, Pulmonalklappenkrankheiten, Kardiomyopathie, Atrioventrikulärer Block, kardiale Erregungsleitungsstörungen, Herzstillstand, Paroxysmale Tachykardie, Vorhofflimmern, kardiale Arrythmie, Herzinsuffizienz. 11) Cerebrovascular diseases. ICD-10: I60, I61, I62, I63, I64, I65, I66, I67, I68, I69. Free-text: Subarachnoidalblutung, Intrazerebrale Blutung, Schlaganfall, Aneurysma, Hämorrhagie. 12) Diseases of arteries. ICD-10: I70, I71, I72, I73, I74, I77, I78, I79. Free-text: Aortenaneurysma, periphere Gefäßkrankheiten, Arterielle Embolie und Thrombose. 13) Diseases of veins. ICD-10: I80, I81, I82, I83, I85, I86, I87, I88, I89. Free-text: Thrombophlebitis, Pfortaderthrombose, sonstige venöse Embolie und Thrombose, Ösophagusvarizen, Varizen, Sonstige Venenkrankheiten, Lymphadenitis, Krankheiten der Lymphgefäße und Lymphknoten. 14) Other and unspecified disorders of the circulatory system. ICD-10: I95, I97, I98, I99. Free-text: Hypotonie. The entries were then inspected and multiple entries per patient for a single disease class/condition were reduced into a single entry, to avoid overrepresentation of a single patient. The number of occurrences of unique patients per disease class/condition were then counted and assembled in a contingency table, with number of patients present for a given disease class/condition and with number of patients absent for a given disease class, for the three groups. Pairwise comparisons were then carried out whereby the data distribution of group I was compared to group II, group I to group III, and group II to group III, for above disease classes/conditions as well as for sex. This resulted in overall 3 x 15 comparisons. Statistical testing was performed using Fisher’s exact test, with significance α at 0.01. P-values ≤ 0.01 were then corrected for multiple comparison using p-value adjustment (Jafari and Ansari-Pour, 2019) where the p-value was multiplied by the number of comparisons performed (i.e. 45). Statistical testing of age was performed using Mann-Whitney U test in GraphPad Prism. Data was visualized with GraphPad Prism as frequencies, i.e. the number of occurrence divided by the total number for each group and each disease class/condition.

#### Mapping the evolution of seroprevalence in two waves according to municipality in the canton of Zurich

The maps of the canton of Zurich were produced using zip code (PLZ) information binned by month for purposes to conserve anonymity. The seroprevalence map (**Fig. 4C** and **D**) displays the ratio of positive versus negative patient samples for each zip code in a given time trace. A threshold of minimally 50 samples per zip code was set in order to minimize statistical fluctuations due to under-sampling a region. This threshold of 50 samples has been made arbitrarily as a tradeoff between the representativity for each zip code and having enough municipalities to represent the sample provenance within the canton of Zurich. In addition, and in order to be able to evaluate any discrepancy, a second map displaying the number of samples analyzed per zip code (independent of seropositivity) has been created (**Fig. 4A** and **B**). The representation of the first six and last six months of the year 2020 has been made in order to compare the evolution of the distribution of seropositivity in the canton of Zurich, between the first and second wave. However, the maps may display significantly lower values than at the seroprevalence peaks as they are averaged over several months. The border of the area of the city of Zurich is surrounded by a dense orange line while the zip codes contained within the canton of Zurich, at the border to another canton, is displayed with a lighter orange line. Limitation of the zip code as representation of the canton of Zurich: A single unique zip code in Switzerland can be shared between several cantons. As the information collected are represented by the zip code, the map generated can partially include municipalities that belong to a canton other than Zurich. These parts are small, however, except for two regions (Baar, Neuhausen am Reinfall). These two regions contain an urban area belonging to cantons other than Zurich (Zug, Schaffhausen respectively) and most likely do not solely represent the Zurich area assigned to this zip code. The following document from the Swiss federal statistical office (Bundesamt für Statistik, 2016) has been used to find the zip code corresponding to the canton of Zurich; the zip codes 5462 and 8363 have been manually added in order to complete the zip code corresponding to the canton of Zurich. The geographical borders corresponding to the zip code border has been taken from the Swiss federal office of topology swisstopo (Bundesamt für Landestopografie, 2020) . We have moreover calculated the averaged seropositivity rate of the city of Zurich and the regions of the Canton not within the city limits binned by 3 months and evaluated the ratio between them.

#### Online health survey

The online health survey was conducted using electronic questionnaires through the REDCap software (https://www.project-redcap.org/). The survey questions are provided here within the codebook (https://github.com/emmarajan/Post_COVID19_online_survey/blob/main/USZ_DIA_PTN_HTA%20Therapeutic%20Antibodies%20(18_0027)%20RED-Cap.pdf); they include questions related to specific symptoms experienced in the 7 days prior to completing the questionnaire, relative health status, the EuroQol 5-dimension 5-level (EQ-5D-5L) and the EuroQol visual analogue scale (EQ VAS) instruments. To calculate EQ-5D-5L health state scores, the value set of the Netherlands was applied in lack of a corresponding value set from Switzerland. Questionnaires could be filled in German or English language. Invitations were sent to potential participants via email by the Clinical Trial Centre at the USZ. All study participants provided electronic consent prior to their participation in the health survey. The implementation of the online health survey was approved by the responsible ethics committee of the canton of Zurich (BASEC-Nr. 2018-01042). Data was collected between 13 April 2022 and 30 May 2022 and a reminder was sent on 03 May 2022 to all participants who had not participated before that date. Data from 142 participants was collected. Data from six individuals was removed from analysis as the survey form was almost entirely incomplete, resulting in a final analysis dataset consisting of 136 individuals. Data was analysed in R 4.2.0 using descriptive statistics and multivariable logistic regression models adjusted for age and sex.

### Protein production

The proteins were produced and purified at different sites in Zurich (CH), Oxford (UK), Lausanne (CH), and Yale University (USA).

#### Oxford, SGC

Recombinant proteins were purified as reported previously with small modifications (Stadlbauer *et al*., 2020; Wrapp *et al*., 2020). Mammalian expression vectors containing secreted, codon-optimized CoV2 S (pHL-Sec (Aricescu, Lu and Jones, 2006); aa. 1-1208, C-terminal 8His- Twin-Strep) and RBD (pOPINTTGNeo; aa. 330-532, C-terminal 6His) were transiently transfected with linear PEI into Expi239^TM^ cells cultured in roller bottles in FreeStyle 293 media. Cell culture media was harvested after 3 days at 37°C for RBD or 3 days at 30°C for Spike and then buffered to 1X PBS. Proteins were first pulled down on Ni^2+^ IMAC Sepharose® 6 Fast Flow (GE) with stringent washing (>50 CV with 40 mM imidazole). RBD was polished on a Superdex 75 16/600 column (GE) equilibrated with 1X PBS, while Spike was directly dialyzed into 1X PBS using SnakeSkin^TM^ 3,500 MWCO dialysis tubing. Proteins were concentrated with VivaSpin® centrifugal concentrators, centrifuged at 21,000 x *g* for 30 min to remove precipitates, and flash frozen at 1 mg/mL

#### Lausanne, EPFL SV PTECH PTPSP and Zurich UZH

The prefusion ectodomain of the CoV2 S protein (the construct was a generous gift from Prof. Jason McLellan, University of Texas, Austin; see (Wrapp *et al*., 2020)) was transiently transfected either into suspension-adapted ExpiCHO cells (Thermo Fisher) or Expi293F (Thermo Fisher) cells with PEI MAX (Polysciences) in ProCHO5 medium (Lonza). After transfection, incubation with agitation was performed at 31°C and 4.5% CO2 for 5 days. The clarified supernatant was purified in two steps; via a Strep-Tactin XT column (IBA Lifesciences) followed by Superose 6 10/300 GL column (GE Healthcare) and finally dialyzed into PBS. The average yield was 15 mg/L culture.

#### Yale, New Haven

Human codon optimized SARS-CoV (2003) RBD (pEZT containing H7 leader sequence; aa. 306-527, C-terminal Aviand 8His tags) was transiently transfected into Expi293^TM^ cells (Thermo Fisher) using the ExpiFectamine^TM^ 293 Transfection kit (Gibco) according to the manufacturer’s instructions. Cells were cultured in a 37°C incubator with 8% humidified CO_2_ for 4 days after transfection. Culture supernatant was collected by centrifugation (500 x g for 10 minutes) and RBD was captured using Ni-NTA Superflow resin (Qiagen), washed, and eluted in buffer containing 50 mM Tris-HCl pH 8, 350 mM NaCl, and 250 mM imidazole. RBD was further purified using an ENrich^TM^ SEC 650 column (Bio-Rad) equilibrated in 1X PBS (Thermo Fisher). Peak fractions were pooled and the protein concentration was determined by 280 nm absorbance with a Nanodrop^TM^ One Spectrophotometer (Thermo Fisher). Protein was snap frozen in liquid nitrogen and shipped on dry ice prior to experiments.

#### Zurich, ETH

NSP1 carrying an N-terminal His6-tag followed by a TEV cleavage site was expressed from a pET24a vector. The plasmid was transformed into E. coli BL21-CodonPlus (DE3)- RIPL and cells were grown in 2xYT medium at 30 °C. At an OD600 of 0.8, cultures were shifted to 18 °C and induced with IPTG to a final concentration of 0.5 mM. After 16 h, cells were harvested by centrifugation, resuspended in lysis buffer (50 mM HEPES-KOH pH 7.6, 500 mM KCl, 5 mM MgCl2, 40 mM imidazole, 10% (w/v) glycerol, 0.5 mM TCEP and protease inhibitors) and lysed using a cell disrupter (Constant Systems Ltd). The lysate was cleared by centrifugation for 45 min at 48.000 xg and loaded onto a HisTrap FF 5-ml column (GE Healthcare). Eluted proteins were incubated with TEV protease at 4 °C overnight and the His6-tag, uncleaved NSP1 and the His6- tagged TEV protease were removed on the HisTrap FF 5-ml column. The sample was further purified via size-exclusion chromatography on a HiLoad 16/60 Superdex75 (GE Healthcare), buffer exchanging the sample to the storage buffer (40 mM HEPES-KOH pH 7.6, 200 mM KCl, 40 mM MgCl2, 10% (w/v) glycerol, 1 mM TCEP). Fractions containing NSP1 were pooled, concentrated in an Amicon Ultra-15 centrifugal filter (10-kDa MW cut-off), flash-frozen in liquid nitrogen, and stored until further use at -80 °C.

#### Details of viral proteins used for this study

For *high-throughput serology*, the following proteins were used: CoV2 S (pHL-Sec; aa. 1-1208, C-terminal 8His-Twin-Strep) and RBD (pOPINTTGNeo; aa. 330-532, C-terminal 6His) produced at the SGC in Oxford and the nucleocapsid protein from AcroBiosystems (AA Met 1 - Ala 419, C-terminal his-tag, NUN-C5227). For *competitive ELISA*, we used: The prefusion ectodomain of the CoV2 S protein (Lausanne, EPFL SV PTECH PTPSP), the RBD from Trenzyme (C-terminal his-tag, P2020-001) and the nucleocapsid protein from AcroBiosystems (AA Met 1 - Ala 419, C-terminal his-tag, NUN-C5227). For *additional ELISAs* following the high-throughput serology, we used: The prefusion ectodomain of the CoV2 S protein (Lausanne, EPFL SV PTECH PTPSP), the RBD from Trenzyme (C-terminal his-tag, P2020-001) and, nucleocapsid protein from AcroBiosystems (AA Met 1 - Ala 419, C-terminal his-tag, NUN-C5227), the COV2 NSP1 protein (from Nenad Ban, ETH Zurich), the CMV pp65 protein (Abcam, ab43041), and BSA (Thermo Scientific).

### Assay validation

#### High-throughput validation screen

For the validation screen, we picked 60 and 150 samples from BDS and USZ, respectively, that had the high average values when summing -logEC_50_ for both Spike and RBD. Additionally, we added 52 and 70 randomly selected prepandemic samples for the BDS and the USZ cohort respectively. We supplemented the three antigens used in the first screen (NC, S, RBD of SARS-COV2) with a SARS-CoV RBD antigen. Unlike for the primary screen, we ran all samples in duplicates spread over two independent plates.

#### Western Blotting

Expi293F cells were obtained as a gift from Prof. Maurizio Scaltriti (Memorial Sloan Kettering Cancer Center, New York). Non transfected control cells and cells overexpressing either His-tagged S, His-tagged NC or His-tagged RBD domain were lysed in 0,1% Triton X- 100/PBS. Total protein content in the cellular fraction was quantified using bicinchoninic protein assay (Pierce BCA Protein Assay Kit, ThermoFisher). For Western Blotting, 30 µg of ECD-expressing lysate, 10 µg of NC-expressing lysate and 10 µg of RBD-expressing lysate were loaded all in the same well of NU-PAGE 4-12% Bis-Tris gels (ThermoFisher). 50 µg of non-transfected cell lysate were loaded as negative control. Gels were run at a constant voltage (150 V) in MES running buffer for 50 minutes, then transferred onto PVDF membrane with a dry transfer system (iBlot 2 Gel Transfer Device, ThermoFisher). The membranes were blocked with 5% SureBlock (Lubio Science) for 1 hour at room temperature, and then incubated overnight with a 1:100 dilution of patients’ plasma in 1% SureBlock, at 4 degrees. The day after, membranes were washed four times with PBS-T and incubated for 1 hours with an anti-human secondary antibody, HRP-conjugated, diluted 1:10000 in 1% SureBlock. The membranes were then washed four times with PBS- T and acquired using Immobilon Crescendo HRP Substrate (Merck Millipore) and Fusion SOLO S imaging system (Vilber). As a positive control, one membrane was incubated overnight with mouse anti-Histag antibody (ThermoFisher, dilution 1:10000 in 1% SureBlock) and subsequently with anti-mouse secondary antibody, HRP-conjugated (Jackson, dilution 1:10000 in 1% SureBlock).

#### 384-well ELISA using multiple antigens

High-binding 384-well plates (Perkin Elmer, SpectraPlate 384 HB) were coated with 20 µL 1 µg/mL S (Lausanne, EPFL SV PTECH PTPSP), RBD (Lausanne, EPFL SV PTECH PTPSP), NC (AcroBiosystems), BSA (ThermoScience), CMV pp65 (abcam, #ab43041), or NSP1 (Zurich, ETH) in PBS at 37 °C for 1 h, followed by 3 washes with PBS 0.1% Tween-20 (PBS-T) using Biotek El406 and by blocking with 40 µL 5% milk in PBS-T for 1.5 h. Serum samples were diluted in sample buffer (1% milk in PBS-T) and a serial dilution (range: 0.005 – 3x10^-7^) was carried out (volume: 20 µL/well). After the sample incubation for 2 h at RT, the wells were washed five times with wash buffer and the presence of IgGs or IgAs directed against above-defined antigens was detected using an HRP-linked anti-human IgG antibody (Peroxidase AffiniPure Goat Anti-Human IgG, Fcγ Fragment Specific, Jackson, 109-035- 098, at 1:4000 dilution in sample buffer) or HRP-linked anti-human IgA antibody (Goat anti-Human IgA (Heavy chain) Secondary Antibody, HRP, 31417, ThermoFisher Scientific, at 1:750 dilution in sample buffer), 20 µL/well. The incubation of the secondary antibody for one hour at RT was followed by three washes with PBS-T, the addition of TMB, an incubation of five minutes at RT, and the addition of 0.5 M H_2_SO_4_. The plates were centrifuged after all dispensing steps, except for the addition of TMB. The absorbance at 450 nm was measured in a plate reader (Perkin Elmer, EnVision) and the inflection points of the sigmoidal binding curves (pEC50 values of the respective sample dilution) were determined using the custom designed fitting algorithm referred to earlier. The pEC50 values for all samples and antigens was visualized using the ggplot2 package in R.

#### Competitive ELISA

To perform competitive ELISAs, high-binding 384-well plates (Perkin Elmer, SpectraPlate 384 HB) were coated with 1 ug/mL S or RBD or NC in PBS at 37 °C for 1 h, followed by 3 washes with PBS-T and by blocking with 5% milk in PBS-T for 1.5 h. Meanwhile, plasma samples were diluted to a final concentration close to the EC_50_, incubated with either RBD (50 ug/mL) or S (12.5 ug/mL) and serially diluted (11 dilution points per patient sample, 25 uL per dilution) in a low-binding 384-well plates (Perkin Elmer, high binding SpectraPlate). After 2 h of incubation at RT, 20 uL of all the samples were transferred to the previously coated plates and incubated for additional 2 h at RT. Then, the plates were washed five times with PBS-T and the presence of IgGs was detected using an HRP-linked anti-human IgG antibody (Peroxidase AffiniPure Goat Anti-Human IgG, Fcγ Fragment Specific, Jackson, 109-035-098, at 1:4000 dilution in sample buffer). The incubation of the secondary antibody for one hour at RT was followed by three washes with PBS-T, the addition of TMB, an incubation of 5 minutes at RT, and the addition of 0.5 M H_2_SO_4_. The absorbance at 450 nm was measured in a plate reader (Perkin Elmer, EnVision). Data were interpreted and the following qualitative categories were assigned: (1) No binding to target protein, no competition. (2) Binding to target protein, no competition. (3) Binding to target protein, competition.

#### Microscale diffusional sizing

For the microfluidic binding measurements, 40% of human plasma was added to 10 nM antigen and PBS was added to give a constant volume of 20 µL. The antigen used was RBD labelled with Alexa Fluor 647 through N-terminal amine coupling. These samples were incubated at room temperature for 40 minutes and the size, hence molecular weight of the formed immunocomplex, was determined through measuring the hydrodynamic radius, Rh with microfluidic diffusional sizing (Arosio *et al*., 2016; Emmenegger, De Cecco, *et al*., 2021; Emmenegger *et al*., 2022; Schneider *et al*., 2022) using a Fluidity One W platform (Fluidic Analytics, Cambridge, UK). Following correction of fluorescence intensities for serum autofluorescence, the fraction, *f_d_*, of RBD to diffuse into the distal channel is defined by:

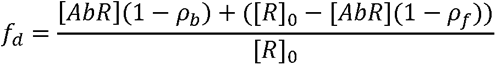

Where [*AbR*] is the concentration of bound RBD, [*R*]_0_ is the total concentration of RBD, and ρ_b_ and ρ_f_ are the fractions of bound and free RBD to diffuse into the distal channel, respectively.

## Data Availability

The raw data underlying this study will be made available upon reasonable request. The biobank samples are limited and were exhausted in several instances. Therefore, while we will make ef-forts to provide microliter amounts of samples to other researchers, their availability is physically limited.

## Acknowledgments

All authors wish to thank their entire teams for support in the lab. Linda Irpinio, André Wethmar, and Andra Chincisan are acknowledged for help in the lab and with data processing. We are grateful to Elisabeth J. Rushing (USZ) for proofreading the manuscript, to Guido Bucklar, Michael Fetzer, Katie Kalt, Karin Edler, Roland Naef, and especially Patrick Hirschi (USZ) for their help with hospital data, and to Didier Trono for helpful insights and discussions. Michael Weisskopf, Antje Thiem, Regina Grossmann and the team of the Clinical Trials Center (CTC) of the USZ are acknowledged for their help with sample acquisition and protocols. Above all, we are grateful to all blood donors and hospital patients for helping us conduct this study. Lastly, Vishalini Emmenegger (ETH Zurich) offered insights, advice and kind help with illustrations, and supported, accompanied, and inspired the work.

## Author contributions

Collected and processed the biological specimens, prepared and carried out the high-throughput screenings, maintained the machines: ME, DS, BD, JG, AW, MI, JD, LB, LM, CZ. Analyzed data from the high-throughput serology: DL, RJ, IX, ME, AA. Carried out follow-up ELISAs and competitive ELISAs: EDC, RM, ME. Carried out Western Blots: EDC, CT, AGG. Carried out and interpreted experiments in solution using microfluidics diffusional sizing technology: MMS, ICM, CKX, GM, TJPK, VK, ME, AA. Collected samples from COVID-19 patients for the establishment of serology: ILD, DJS. Coordinated the sample acquisition and processing from the Institute of Clinical Chemistry: AvE and LS. Coordinated the sample acquisition and processing from the blood donation services: BF and JG. Coordinated and performed high-throughput ELISAs for comparison in Oxford: DE, StH, DIS. Coordinated and performed CoV2 serological assays using commercial platforms: LS, KS, AvE, EPM, OB. Produced proteins: DIS, NBB, RO, TM, FP, DH, KL, EDC, JDH, FL, AMR, SH, GS. Carried out the epidemiological modeling: CA, JR, ME. Designed, conducted, analyzed, and interpreted the online health questionnaire: TB, DM, MAP, ME. Coordinated the correspondence with the ethics committee of the Kanton of Zurich: RR, JN, ME. Produced and redacted the figures: AA, RJ, DL, EDC, ME. Conceived the idea of creating a biobank of plasma samples, supervised the study on a daily basis, proposed primary and confirmatory experiments, advised on best lab practices, on control experiments and on data interpretation: AA. Wrote abstract, introduction and discussion: AA. Wrote a first draft of the Results part of the manuscript: ME. Advised on and corrected the Results section: AA. All authors contributed by reviewing the first draft, approved the final version, and consented to be accountable for the work.

## Data and materials availability

The raw data underlying this study will be made available upon reasonable request. The biobank samples are limited and were exhausted in several instances. Therefore, while we will make efforts to provide microliter amounts of samples to other researchers, their availability is physically limited.

## Supplementary Figures

**Fig. S1.**
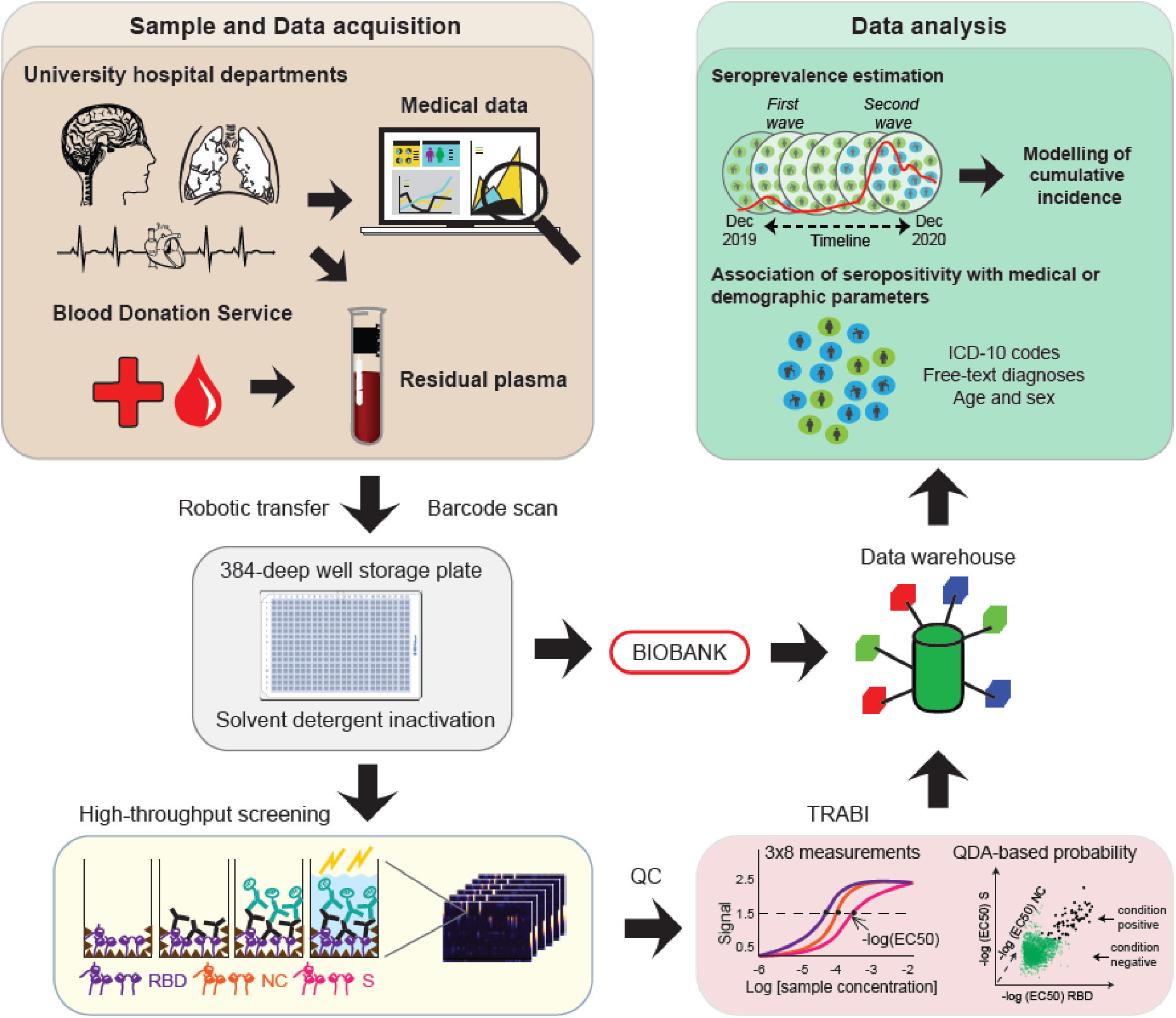
Detailed assay procedure. Barcoded residual plasma samples from patients admitted to almost all clinical departments of the University Hospital Zurich (USZ) or from healthy donors donating blood to the Blood Donation Services (BDS) Canton Zurich were robotically transferred into 384-deep well plates and subjected to viral inactivation using a solvent detergent procedure. The plates were subsequently stored in a liquid high-throughput biobank, while associated medical data were stored in a custom-designed data warehouse. The samples were used to conduct high-throughput serological assays to interrogate for the presence of antibodies against multiple CoV2 proteins, with an indirect ELISA. To this end, plates were coated with S, RBD, or NC. Following the mapping of ELISA results, antibody titers (i.e. -log(EC_50_)) against the three antigens were determined from eight sample dilutions via logistic regression. The -log(EC_50_) profiles from both condition negatives (prepandemic samples) and condition positives (convalescent CoV2- PCR-confirmed individuals for BDS and COVID-19 patients for USZ) were used to infer the QDA-based posterior probability of each sample to be seropositive and data was stored in afore mentioned data warehouse. The data was then used for seroprevalence estimation and the modelling of the cumulative incidence. Medical and demographic data was used to interrogate the presence of potential associations between disease and CoV2 positivity.

**Fig. S2.**
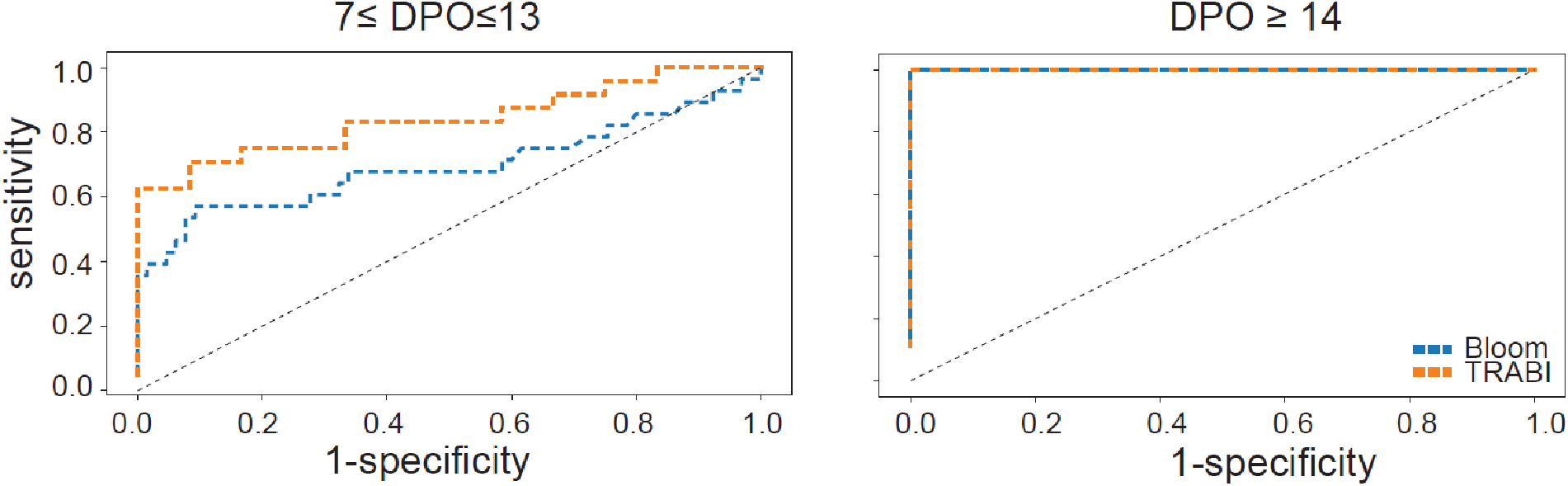
Comparison of TRABI to a lateral-flow assay developed by Bloom diagnostics.

**Fig. S3.**
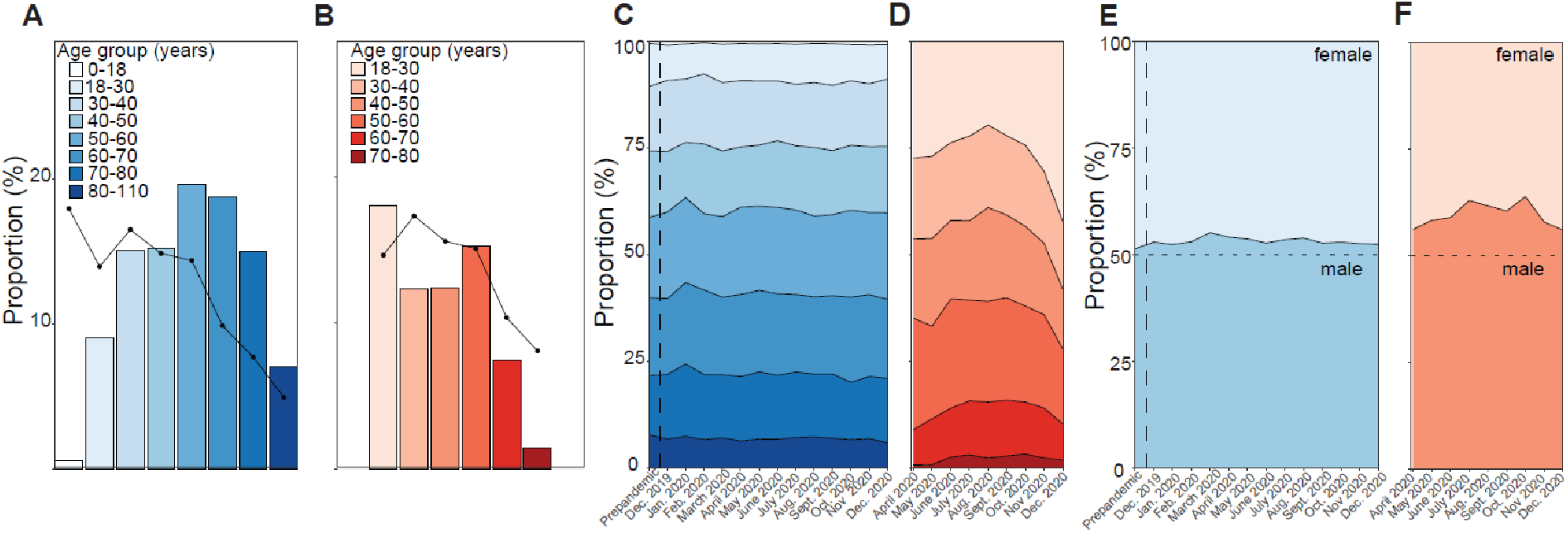
Age and sex characteristics of the two cohorts. **A** and **B**. Sample distribution according to age groups for USZ (A) and for BDS (B). **C** and **D**. Proportion of age groups over time for USZ (C) and for BDS (D). **E** and **F**. Sex distribution over time for USZ (E) and for BDS (F).

**Fig. S4.**
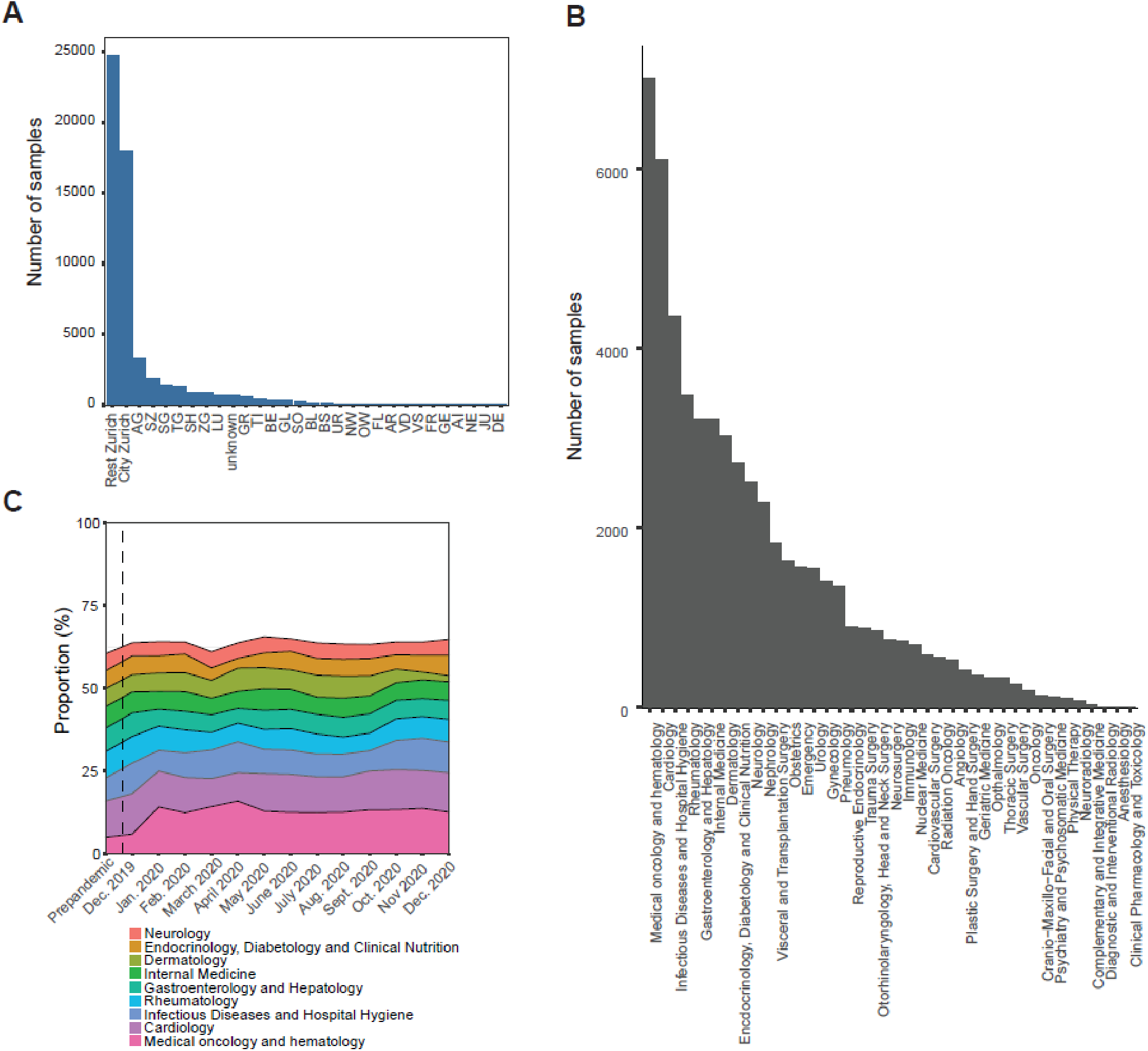
Sample provenance and clinical wards of the USZ patients. **A**. Sample origin for USZ patients. **B**. Clinical departments at which patients were treated. **C**. Distribution of clinical departments over time of study.

**Fig. S5.**
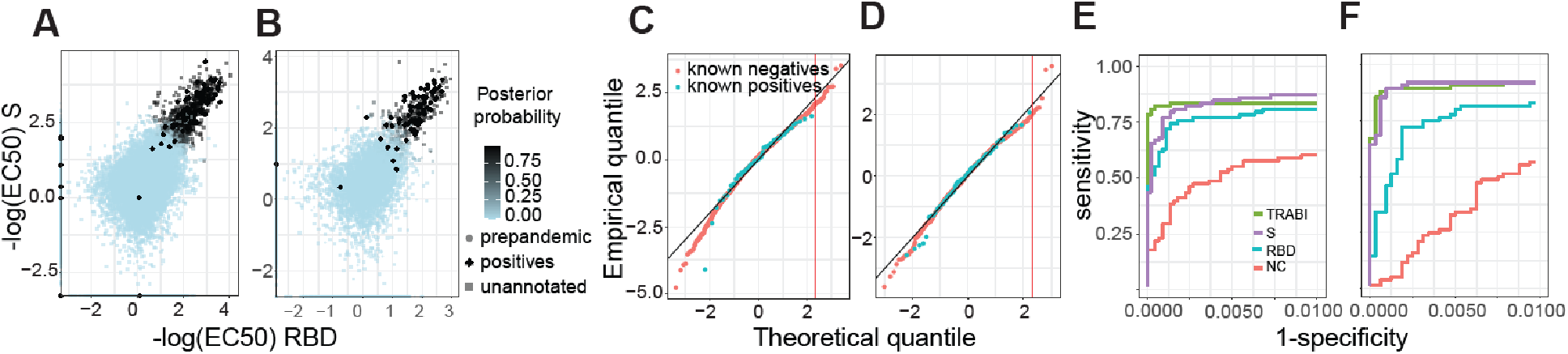
Prevalence estimation in two large cohorts. **A.** and **B**. Depicted are all the –log(EC_50_) values calculated for S and the RBD for the USZ (A) and the BDS (B) cohort. Posterior probability were calculated using LDA. **C**. and. **D**. Q-Q plots to check the Gaussian distributional assumption of the LDA model. For USZ (C) and the BDS (D) cohort respectively, we collapsed the 3 measures per sample according to the linear discriminant classifier. We then scaled known positives and negatives to mean zero and unit variance and compared their distributions to the univariate Gaussian distribution via Q-Q plot. We saw that for the relevant upper tail, the distribution of known negatives followed the normal distribution with only a mild deviation, such that, for instance, the upper one percent quantile is reached at 2.07 (C) and 1.99 (D) rather than at their theoretical value of 2.33. **E**. and. **F**. ROC curves for the USZ (E) and BDS (F) cohorts using the prepandemic samples (including condition negatives from December 2019 and January 2020 for BDS) as condition negatives and selected condition positives from both cohorts.

**Fig. S6.**
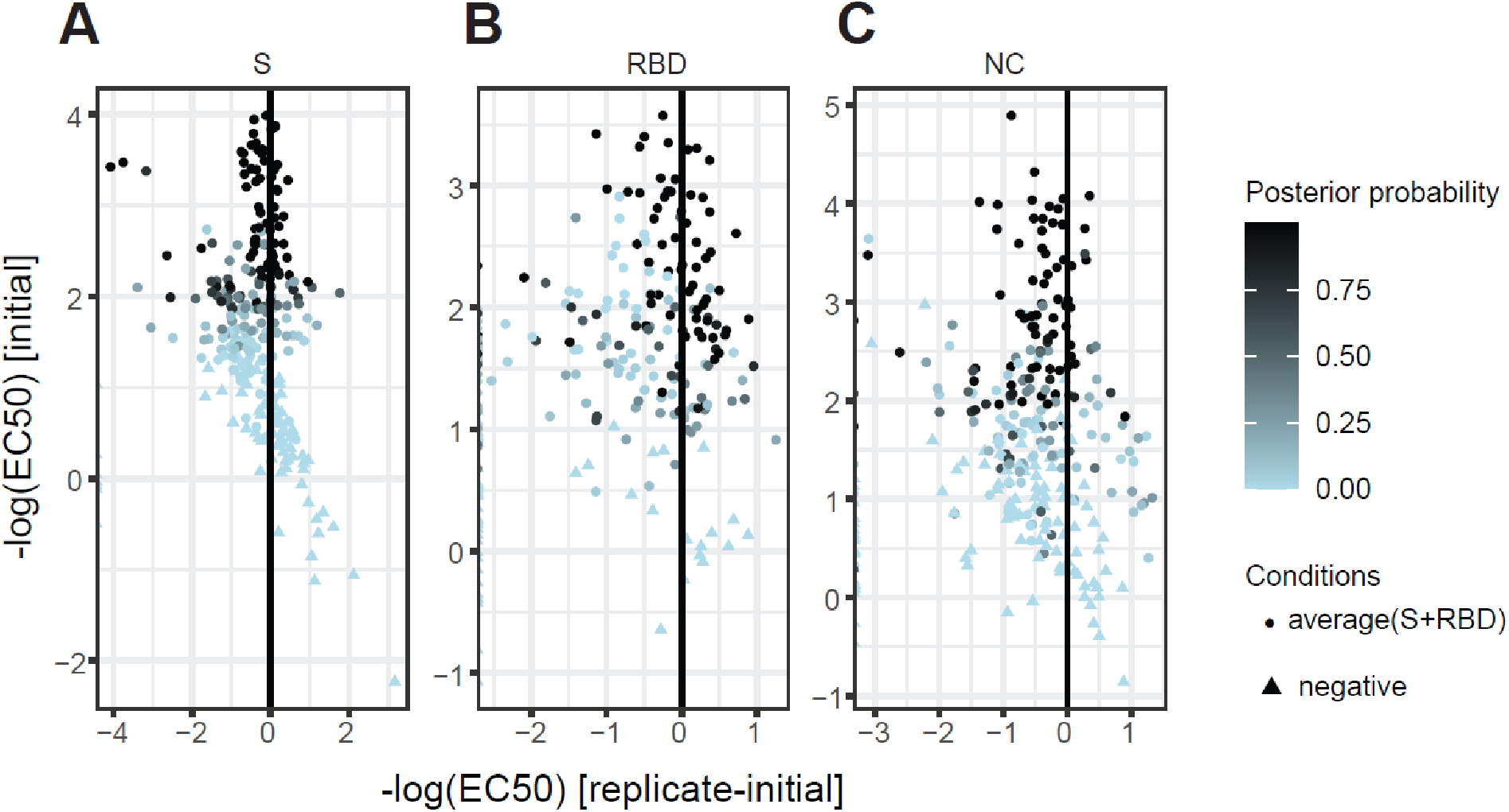
Assay reproducibility using 210 high scoring samples and 122 random samples (based on results from the high-throughput screen) for binding against S, the RBD, and the NC. **A.** S binding shows that reproducibility increases at higher values, consistent with increased posterior probabilities. **B.** Same observation as (A) for the RBD. **C**. Same observation as (A) for the NC.

**Fig. S7.**
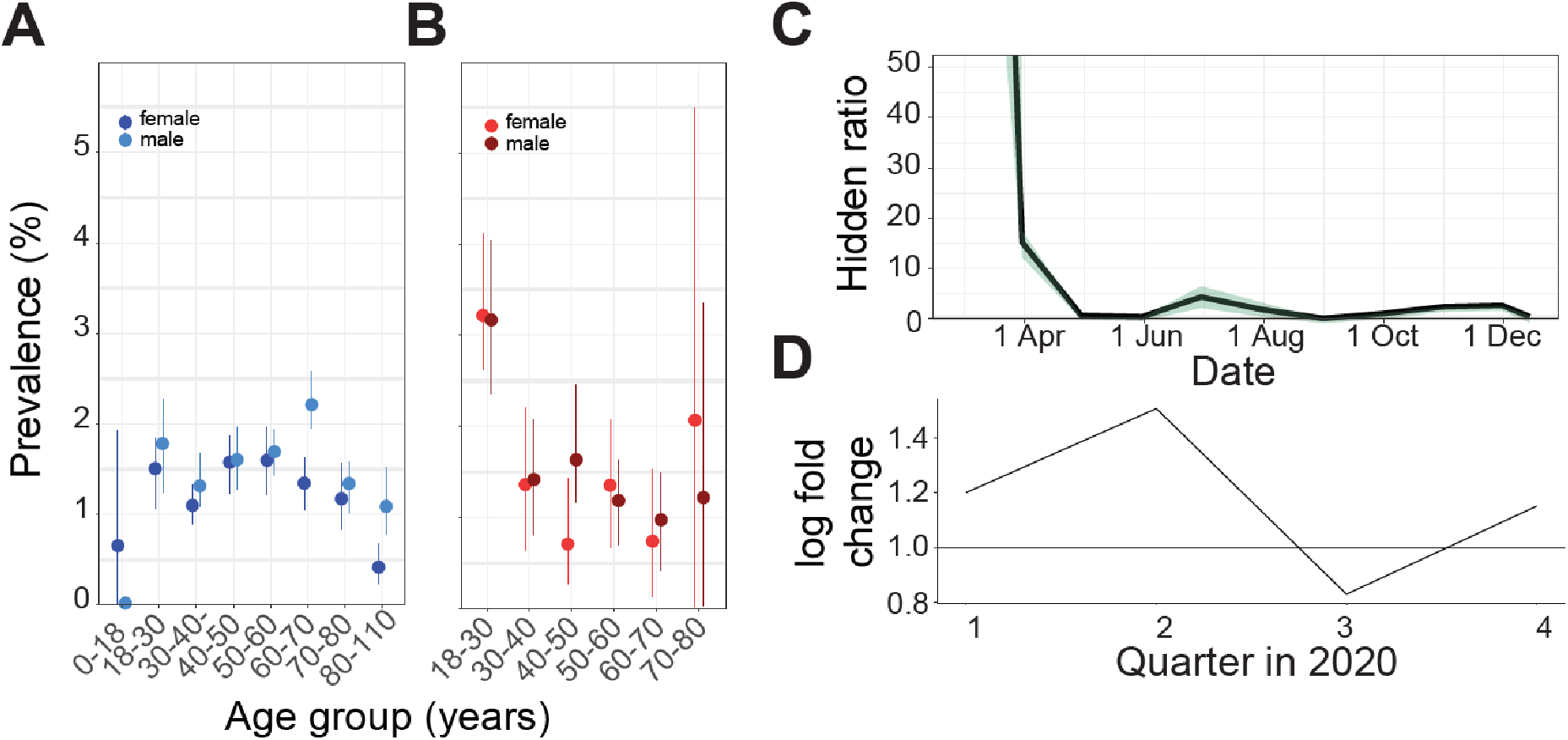
Epidemiological modelling. **A**. Seroprevalence according to age and sex for the hospital patients. **B**. Seroprevalence according to age and sex for the blood donors. **C**. Hidden epidemic ratio. **D**. Log fold change of seropositivity of hospital patients from the city versus non-city in four quarters, covering the first and the second wave.

**Fig. S8.**
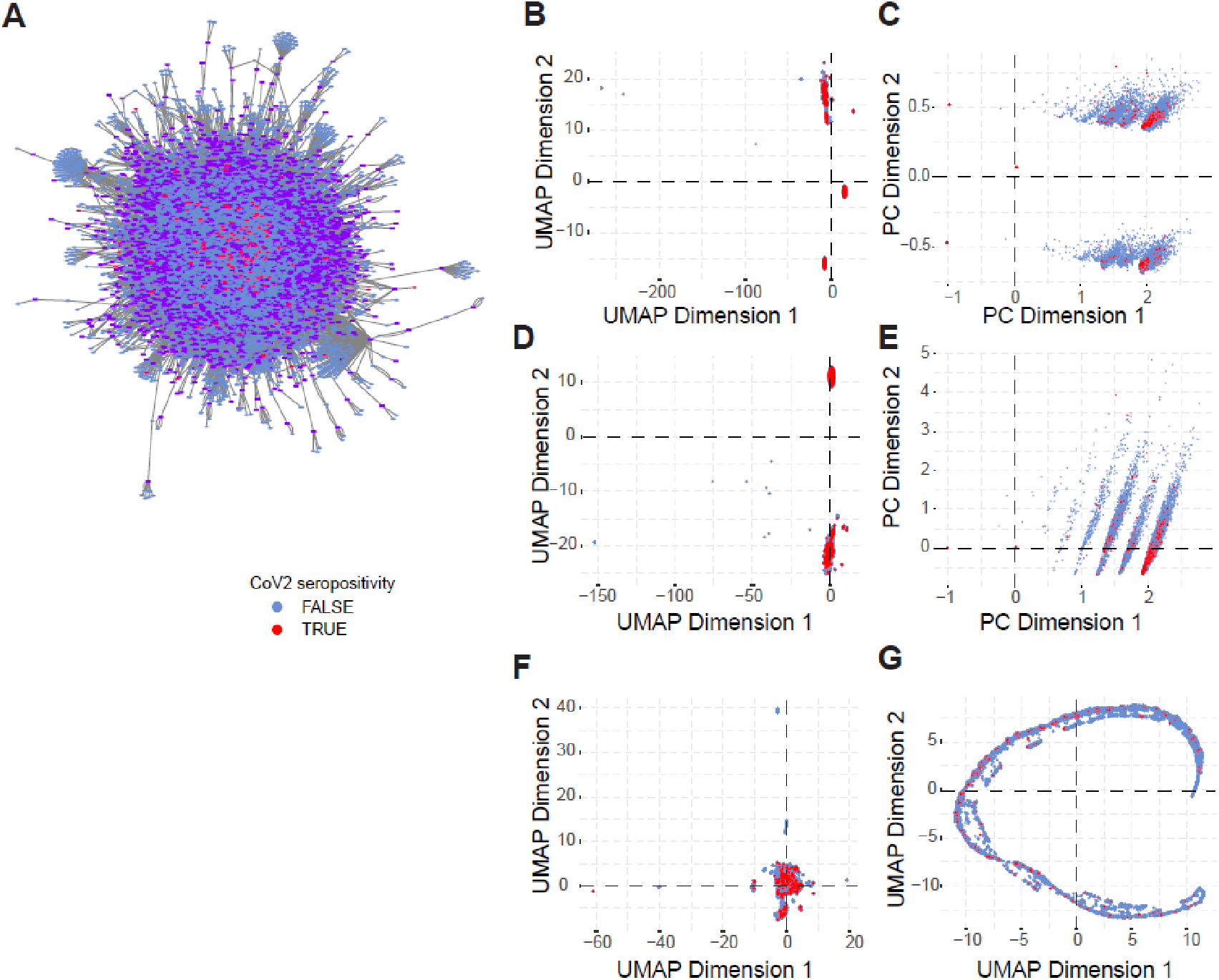
Exploratory analysis of CoV2 seropositivity with ICD-10 codes and free-text medical reports. **A**. Force directed layout of topological network. ICD-10 codes: purple rectangles. Male patients: diamonds, female patients: circles. Seropositive: red. Seronegative: blue. **B**. Two-dimensional projections of ICD-10 codes as well as female and male sex following UMAP tuned for binary data input using cosine metric. **C**. Principal component analysis of ICD-10 codes as well as female and male sex following. **D**. and **E**. Same as in (B) and (C) with exclusion of male/female sex as feature. **F**. and **G**. Representation of dataset with exclusion of patients for which no ICD-10 data exists, using cosine (F) or Euclidean (G) distances. For A-G, seropositivity: red. Seronegativity: blue. No distributional differences between seropositive and seronegative hospital patients were visible.

**Fig. S9.**
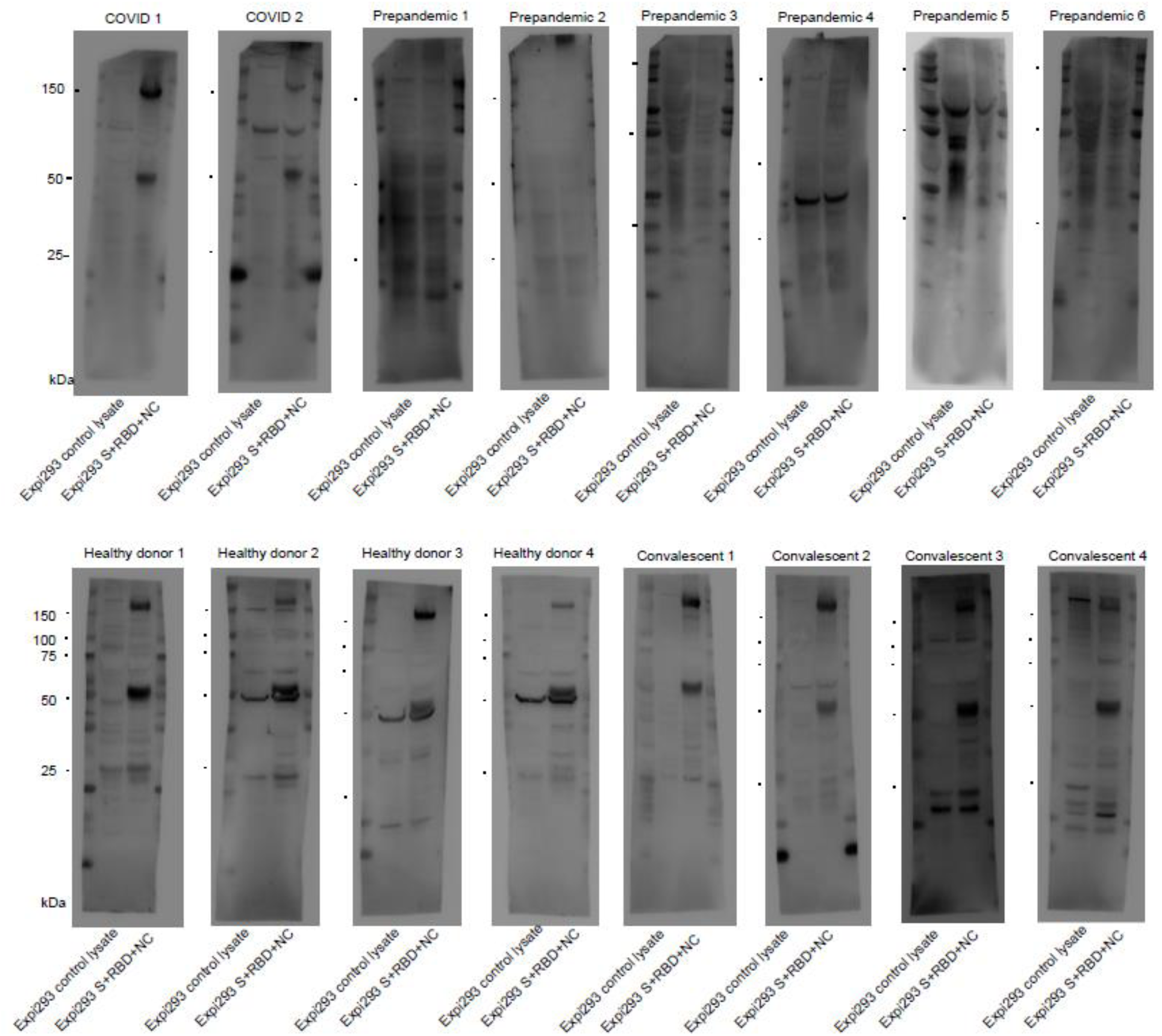
Uncropped and unmodified camera-acquired images of the Western Blots.

**Table S1.** Characterization of cohort used for assay establishment.

|  | Total | No oxygen supply | Oxygen supply | Intubation |
| --- | --- | --- | --- | --- |
| Individuals, number | 27 | 8 | 12 | 7 |

**Table S2.** 50 most common ICD-10 codes for hospital patients.

| ICD-10 | ICD-10 Text | Nr. of patients with code |
| --- | --- | --- |
| I10.00 | Essentielle (primäre) Hypertonie | 3251 |
| U99.0 | Spezielle Verfahren zur Untersuchung auf SARS-CoV-2 | 2790 |
| Z11 | Spezielle Verfahren zur Untersuchung auf infektiöse und parasitäre Krankheiten | 2773 |
| Y84.9 | Zwischenfälle durch medizinische Maßnahmen, nicht näher bezeichnet | 1756 |
| Z92.1 | Dauertherapie (gegenwärtig) mit Antikoagulanzen in der Eigenanamnese | 1615 |
| N18.3 | Chronische Nierenkrankheit, Stadium 3 | 1259 |
| I48.0 | Vorhofflimmern, paroxysmal | 1218 |
| D90 | Immunkompromittierung nach Bestrahlung, Chemotherapie und sonstigen immunsuppressiven Maßnahmen | 1184 |
| U69.12 | Temporäre Blutgerinnungsstörung | 1096 |
| B96.2 | Escherichia coli [E. coli] und andere Enterobacterales als Ursache von Krankheiten, die in anderen Kapiteln klassifiziert sind | 1072 |
| Z86.7 | Krankheiten des Kreislaufsystems in der Eigenanamnese | 1001 |
| Z95.5 | Vorhandensein eines Implantates oder Transplantates nach koronarer Gefäßplastik | 992 |
| E11.90 | Diabetes mellitus, Typ 2, mit Koma | 904 |
| I25.13 | Drei-Gefäß-Erkrankung | 828 |
| D69.58 | Sonstige sekundäre Thrombozytopenien, nicht als transfusionsrefraktär bezeichnet | 820 |
| E55.9 | Vitamin-D-Mangel, nicht näher bezeichnet | 816 |
| E78.4 | Sonstige Hyperlipidämien | 795 |
| D62 | Akute Blutungsanämie | 793 |
| Y82.8 | Zwischenfälle durch medizintechnische Geräte und Produkte | 776 |
| E87.6 | Hypokaliämie | 734 |
| F05.8 | Sonstige Formen des Delirs | 722 |
| Y57.9 | Komplikationen durch Arzneimittel oder Drogen | 711 |
| E03.8 | Sonstige näher bezeichnete Hypothyreose | 673 |
| I50.01 | Rechtsherzinsuffizienz, Sekundäre Rechtsherzinsuffizienz | 656 |
| Z92.2 | Dauertherapie (gegenwärtig) mit anderen Arzneimitteln in der Eigenanamnese | 653 |
| J96.00 | Akute respiratorische Insuffizienz, anderenorts nicht klassifiziert | 638 |
| I50.12 | Linksherzinsuffizienz, Mit Beschwerden bei stärkerer Belastung | 629 |
| I25.11 | Ein-Gefäß-Erkrankung | 619 |
| N39.0 | Harnwegsinfektion, Lokalisation nicht näher bezeichnet | 614 |
| I25.12 | Zwei-Gefäß-Erkrankung | 606 |
| X59.9 | Sonstiger und nicht näher bezeichneter Unfall | 597 |
| D68.4 | Erworbener Mangel an Gerinnungsfaktoren | 589 |
| N40 | Prostatahyperplasie | 570 |
| I10.90 | Essentielle Hypertonie, nicht näher bezeichnet | 566 |
| C43.9 | Bösartiges Melanom der Haut, nicht näher bezeichnet | 551 |
| E78.8 | Sonstige Störungen des Lipoproteinstoffwechsels | 547 |
| E87.1 | Hypoosmolalität und Hyponatriämie | 531 |
| I34.0 | Mitralklappeninsuffizienz | 526 |
| I11.90 | Hypertensive Herzkrankheit ohne (kongestive) Herzinsuffizienz | 507 |
| Z95.88 | Vorhandensein von sonstigen kardialen oder vaskulären Implantaten oder Transplantaten | 504 |
| E11.91 | Diabetes mellitus, Typ 2, mit Ketoazidose | 493 |
| E83.38 | Sonstige Störungen des Phosphorstoffwechsels und der Phosphatase | 493 |
| U69.00 | Anderenorts klassifizierte, im Krankenhaus erworbene Pneumonie | 491 |
| I50.14 | Linksherzinsuffizienz, Mit Beschwerden in Ruhe | 476 |
| I50.13 | Linksherzinsuffizienz, Mit Beschwerden bei leichter Belastung | 472 |
| D50.8 | Sonstige Eisenmangelanämien | 467 |
| E78.0 | Reine Hypercholesterinämie | 455 |
| D64.8 | Sonstige näher bezeichnete Anämien | 445 |
| J91 | Pleuraerguss bei anderenorts klassifizierten Krankheiten | 443 |
| A49.8 | Sonstige bakterielle Infektionen nicht näher bezeichneter Lokalisation | 438 |

**Table S3.**
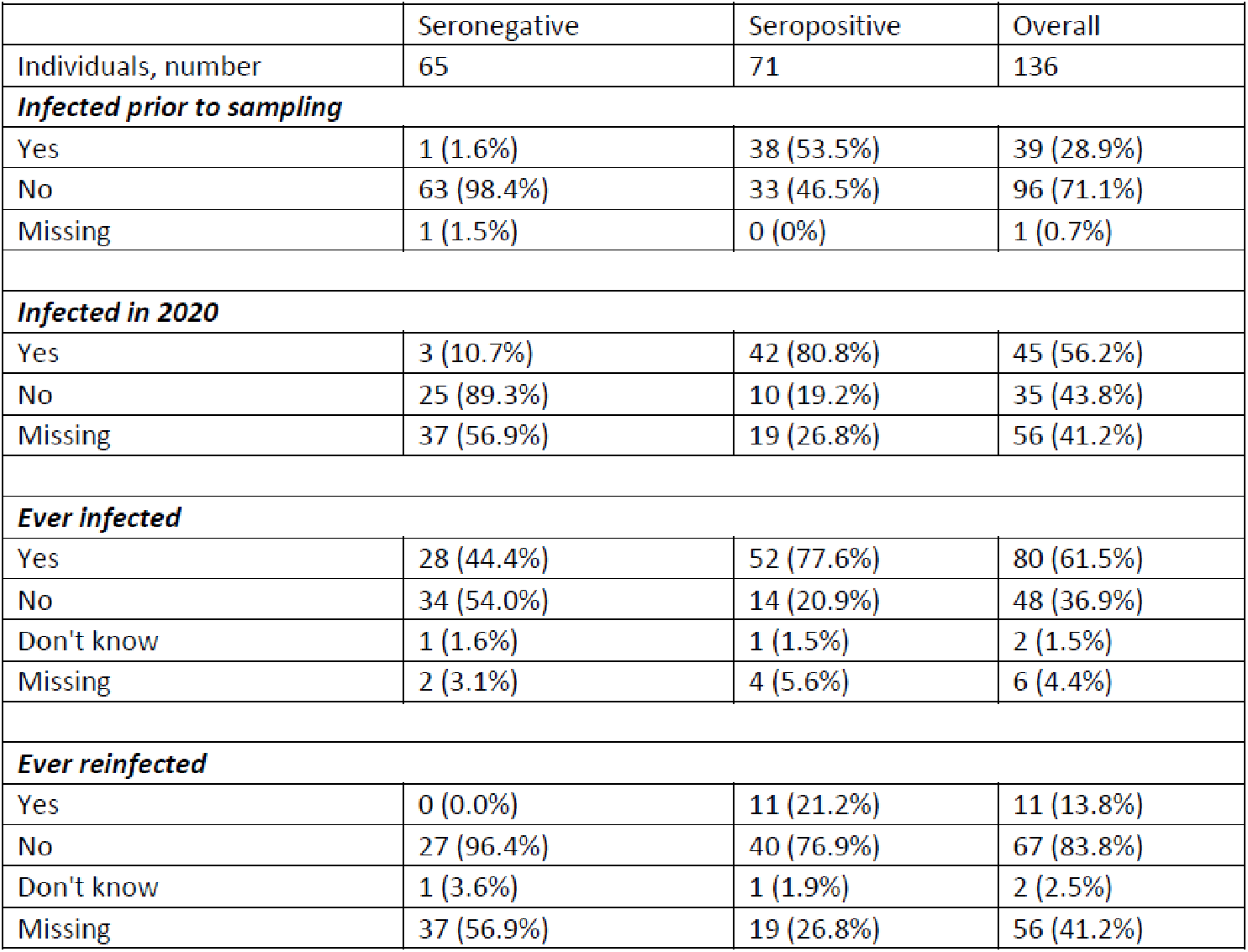
Knowledge of infection prior to blood sampling among online health survey participants tested seropositive or seronegative.

|  | Seronegative | Seropositive | Overall |
| --- | --- | --- | --- |
| Individuals, number | 65 | 71 | 136 |
| ***Infected prior to sampling*** |  |  |  |
| Yes | 1 (1.6%) | 38 (53.5%) | 39 (28.9%) |
| No | 63 (98.4%) | 33 (46.5%) | 96 (71.1%) |
| Missing | 1 (1.5%) | 0 (0%) | 1 (0.7%) |
| ***Infected in 2020*** |  |  |  |
| Yes | 3 (10.7%) | 42 (80.8%) | 45 (56.2%) |
| No | 25 (89.3%) | 10 (19.2%) | 35 (43.8%) |
| Missing | 37 (56.9%) | 19 (26.8%) | 56 (41.2%) |
| ***Ever infected*** |  |  |  |
| Yes | 28 (44.4%) | 52 (77.6%) | 80 (61.5%) |
| No | 34 (54.0%) | 14 (20.9%) | 48 (36.9%) |
| Don't know | 1 (1.6%) | 1 (1.5%) | 2 (1.5%) |
| Missing | 2 (3.1%) | 4 (5.6%) | 6 (4.4%) |
| ***Ever reinfected*** |  |  |  |
| Yes | 0 (0.0%) | 11 (21.2%) | 11 (13.8%) |
| No | 27 (96.4%) | 40 (76.9%) | 67 (83.8%) |
| Don't know | 1 (3.6%) | 1 (1.9%) | 2 (2.5%) |
| Missing | 37 (56.9%) | 19 (26.8%) | 56 (41.2%) |

**Table S4.** Severity of infection among online health survey participants reporting a known CoV2 infection up to April/May 2022, stratified by infection wave.

|  | Spring/Summer<br>2020 | Fall/Winter<br>2020/2021 | Winter/Spring<br>2021/2022 | Overall |
| --- | --- | --- | --- | --- |
| Individuals,<br>number | 12 | 34 | 34 | 80 |
| **Symptoms at infection** |  |  |  |  |
| Yes | 10 (90.9%) | 25 (73.5%) | 29 (85.3%) | 64 (81.0%) |
| No | 1 (9.1%) | 9 (26.5%) | 5 (14.7%) | 15 (19.0%) |
| Missing | 1 (8.3%) | 0 (0%) | 0 (0%) | 1 (1.3%) |
| **Hospitalisation due to COVID-19** |  |  |  |  |
| Yes | 5 (41.7%) | 8 (23.5%) | 1 (2.9%) | 14 (17.5%) |
| No | 7 (58.3%) | 26 (76.5%) | 33 (97.1%) | 66 (82.5%) |
| **Oxygen supplementation** |  |  |  |  |
| Yes | 4 (80.0%) | 6 (75.0%) | 1 (100.0%) | 11 (78.6%) |
| No | 1 (20.0%) | 2 (25.0%) | 0 (0.0%) | 3 (21.4%) |
| Missing | 7 (58.3%) | 26 (76.5%) | 33 (97.1%) | 66 (82.5%) |
| **Intensive care treatment** |  |  |  |  |
| Yes | 4 (80.0%) | 3 (37.5%) | 1 (100.0%) | 8 (57.1%) |
| No | 1 (20.0%) | 5 (62.5%) | 0 (0.0%) | 6 (42.9%) |
| Missing | 7 (58.3%) | 26 (76.5%) | 33 (97.1%) | 66 (82.5%) |
| **Mechanical ventilation** |  |  |  |  |
| Yes | 2 (50.0%) | 3 (100.0%) | 1 (100.0%) | 6 (75.0%) |
| No | 2 (50.0%) | 0 (0.0%) | 0 (0.0%) | 2 (25.0%) |
| Missing | 8 (66.7%) | 31 (91.2%) | 33 (97.1%) | 72 (90.0%) |
| **Diagnosed pneumonia** |  |  |  |  |
| Yes | 3 (25.0%) | 5 (15.2%) | 1 (3.0%) | 9 (11.5%) |
| No | 8 (66.7%) | 28 (84.8%) | 29 (87.9%) | 65 (83.3%) |
| Don't know | 1 (8.3%) | 0 (0.0%) | 3 (9.1%) | 4 (5.1%) |
| Missing | 0 (0%) | 1 (2.9%) | 1 (2.9%) | 2 (2.5%) |

**Table S5.** Health status measured by EQ-5D-5L and EQ VAS among online health survey participants reporting a known CoV2 infection up to April/May 2022, stratified by infection wave.

|  | Never infected | Infected | Overall |
| --- | --- | --- | --- |
| Individuals, number | 50 | 80 | 136 |
| **EQ-5D-5L score** |  |  |  |
| Mean (SD) | 0.81 (0.17) | 0.87 (0.19) | 0.85 (0.18) |
| Median (IQR) | 0.85 (0.74 to 1.00) | 0.91 (0.83 to 1.00) | 0.89 (0.79 to 1.00) |
| Range | 0.42 to 1.00 | -0.02 to 1.00 | -0.02 to 1.00 |
| Missing | 5 (10.0%) | 11 (13.8%) | 20 (14.7%) |
| **EQ VAS score** |  |  |  |
| Mean (SD) | 70.30 (20.88) | 75.00 (15.83) | 72.92 (18.18) |
| Median (IQR) | 74.50 (60.75 to 89.50) | 78.50 (70.25 to 84.75) | 76.00 (70.00 to 85.00) |
| Range | 18.00 to 97.00 | 10.00 to 100.00 | 10.00 to 100.00 |
| Missing | 4 (8.0%) | 10 (12.5%) | 18 (13.2%) |

